# Placental DNA methylation captures shared and trait-specific genetic susceptibility across complex health conditions

**DOI:** 10.64898/2026.07.07.26357471

**Authors:** Ariadna Cilleros-Portet, Itziar González-Moro, Hachem Sadikki, Sergi Marí, Todd M. Everson, Alba Hernangómez-Laderas, Lucile Broséus, Jorg Tost, Marta Cosín-Tomàs, Marika Groleau, Darina Czamara, Manuel Lozano, Ke Hao, Johanna Tuhkanen, M. Daniele Fallin, Rebecca J. Schmidt, Charles E. Breeze, J.F. Deleuze, Sofia Aguilar-Lacasaña, Pierre-Etienne Jacques, Soile Hytti, Amaia Irizar, Marius Lahti-Pulkkinen, Kelly M. Bakulski, John Dou, Jari Lahti, Martine Vrijheid, Katri Räikkönen, Marie-France Hivert, Jordi Sunyer, Barbara Heude, Johanna Lepeule, Stephanie J. London, Jia Chen, Mariona Bustamante, Carmen J. Marsit, Jose Ramon Bilbao, Corina Lesseur, Nora Fernandez-Jimenez

## Abstract

The Developmental Origins of Health and Disease (DOHaD) hypothesis proposes that the perinatal environment shapes susceptibility to complex traits across life [1]. The placenta, a transient organ mediating maternal-fetal exchange, plays a central role in this process and has emerged as a key molecular archive *in utero* [2–4]. Placental DNA methylation (DNAm) is a unique mediator between prenatal exposures, fetal genetics and later-life outcomes [5–9]. DNAm quantitative trait loci (mQTL) have helped disentangling causal mechanisms underlying GWAS loci for complex diseases [10–15]. Despite growing evidence that placental genomic regulation has broad and profound effects on the developmental programming of early– and later-life health outcomes [17], existing placental studies remain limited in scale and largely focused on growth-and neuro-related traits [12–16]. Here, we construct a high-resolution placental mQTL resource and systematically investigate how placental DNAm relates to early– and later-life traits, and to shared vulnerability and complex interactions among them.

**Teaser:** Placental DNA methylation reveals molecular clues to how life before birth shapes health across the lifespan.

## Main

To investigate how genetically regulated placental DNA methylation (DNAm) relates to complex trait susceptibility, we constructed a placental methylation quantitative trait loci (mQTL) resource comprising 2,583 placentas from nine international cohorts of the Pregnancy And Childhood Epigenetics (PACE) consortium. After stringent quality control and harmonization, we identified widespread genetic regulation of placental DNAm across the genome (Fig. 1; Supplementary Figs. 1-4; Supplementary Note 1).

**Figure 1.**
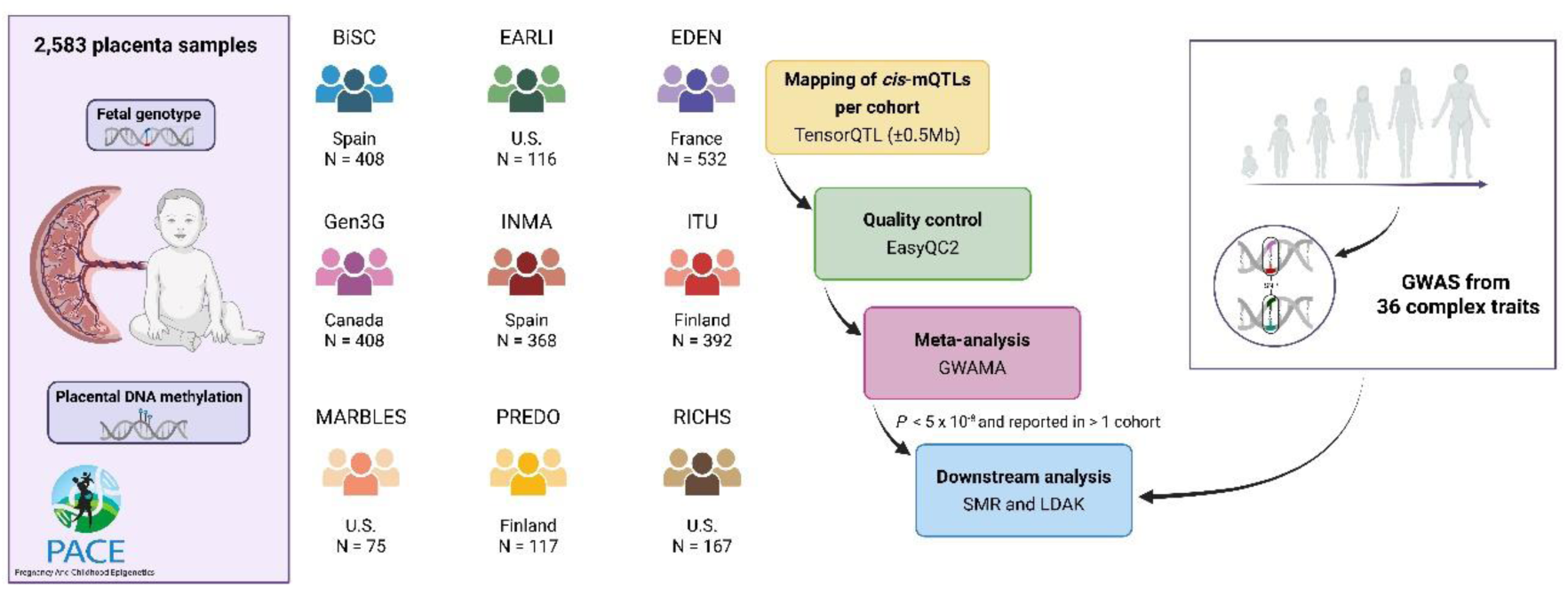
Diagram of the study flowchart, including the list of the PACE cohorts participating in the study. See Methods and Supplementary Note for cohort-specific details. BiSC, EDEN (EPIC), Gen3G, INMA, ITU, MARBLES and PREDO were analyzed with the Illumina EPIC array (N = 1,971). EDEN (450k), EARLI and RICHS were analyzed with the Illumina 450K array (N = 612). All probes were considered for the meta-analysis. More details regarding the cohorts can be found in Supplementary Data 1.

We performed *cis*-mQTL analyses within each cohort using linear regression and a *cis*-window of ±0.5 Mb around each CpG site. Genomic inflation (λ) of cohort-level results ranged from 1.07 to 1.3 (Supplementary Data 1). Next, we performed an inverse variance weighted fixed-effects meta-analysis using GWAMA [18]. We discovered more than 44 million *cis*-mQTLs corresponding to 289,101 CpGs (mSites) with at least one SNP (mVariant) (*P*_meta_ < 5×10^−8^) and significant associations reported in more than one cohort under a cohort-specific nominal P < 5×10^−8^. Most mQTLs (76.8%) were reported in at least half of the cohorts (median of 6) and in a median of 1,896 samples (Supplementary Fig. 1A). Most meta-analysis I^2^ heterogeneity estimates clustered around 0, indicating homogeneous mQTL effects across studies (Supplementary Fig. 1B). mVariants were located at an absolute median distance of 66.23 kb (interquartile range = 129.67) from the mSite (Supplementary Fig. 1C). In addition to the primary placental mQTL database, we also defined a lead mVariant database with the top SNP for each mSite. These databases and complete summary statistics are publicly available at: https://pace-placenta-mqtl.streamlit.app/. This represents the largest placental mQTL catalogue, as well as the largest generated in any tissue comprising the Illumina EPIC probes.

Placental mSites were enriched in CpG island-distal and intermediate DNAm regions, as well as in placenta-specific accessible chromatin and transcriptionally active regions (Supplementary Figs. 2D-E, 3). mSites were enriched for seven transcription factor binding sites (GR, FOS, GATA3, IKZF1, p300, FOXA1, and ESR1) (Supplementary Fig. 2C and Supplementary Data 4). Analyses with GO and KEGG gene sets showed enrichment for cell proliferation and differentiation, as well as for metabolic and immune-related pathways (Supplementary Fig. 4).

We assessed pleiotropic associations with the multi-SNP-based Mendelian Randomization (MR) test of the Summary-based MR (SMR), between placental *cis*-mQTLs and 36 early– and later-life complex traits spanning anthropometric, neurological, metabolic, cardiovascular, digestive and immune domains (Fig. 2 and Supplementary Data 6). Fig. 2A displays the number of placental mSites that are a significant SMR hit, grouped by trait categories (Bonferroni P_SMR_ < 0.05 and P_HEIDI_ > 0.05) (Supplementary Data 7, 8). Overall, the body measurements category exhibited the greatest total number of significant hits (N = 20,149); body height had the highest number of significant CpG sites with 12,339 hits, followed by body mass index (BMI) (N = 4,182) and waist-to-hip ratio (WHR) (N = 2,652). Schizophrenia (SCZ) and hypertension were also prominent with 1,580 and 1,571 pleiotropic CpG sites, respectively (Fig. 2A). We compared placental mSites that were significant SMR hits with those obtained in a systemic tissue (Supplementary Data 9), the whole blood mQTL database from Hannon *et al.* 2018 [19] (N = 1,175, European ancestry, EPIC array). Most traits had a higher number of SMR hits in placenta than in blood (Fig. 2A and Supplementary Data 8). Furthermore, placenta hits tended to be exclusively detected in this tissue (Supplementary Fig. 5 and Supplementary Data 8).

**Figure 2.**
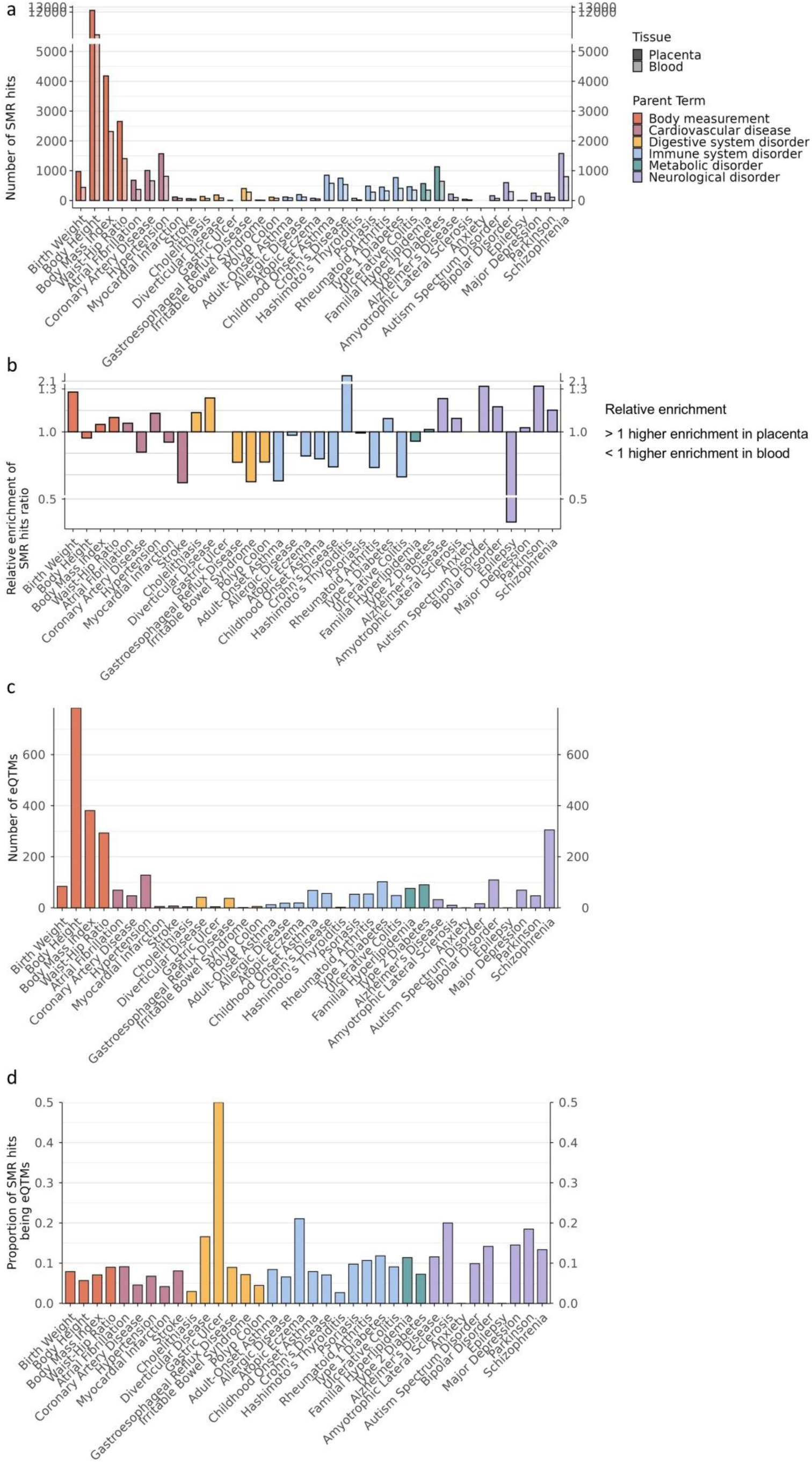
SMR and eQTMs results from the 36 complex traits analyzed. In all the plots, the X-axis represents each of the complex traits, and each of them has been color-coded as a function of its parent term. In plot a) the number of SMR hits obtained per trait is represented in the Y-axis. The darker bar represents placenta and the lighter one whole blood. b) Depicts the relative enrichment of SMR hits in placenta versus whole blood. The proportion for each tissue/fluid was calculated by considering the significant hits obtained in each of the traits out of the total number of CpG sites tested. The enrichment score was computed by dividing the proportion in placenta by the proportion obtained in whole blood. Considering this, a value > 1 suggests greater relative enrichment in placenta, while a value < 1 indicates higher enrichment in whole blood. Plot c) shows the number of placental SMR hits that are also placental eQTMs, while plot d) shows their proportion relative to the total SMR hits per trait in their corresponding Y-axes. The data used for these plots can be found in Supplementary Data 8.

To avoid bias from sample size and power differences between mQTL databases, we computed the enrichment ratio for each trait (Fig. 2B) by dividing the proportion of trait-associated hits found among all tested CpGs in placenta, by the same proportion in whole blood (values > 1 indicate higher enrichment in the placenta) (Supplementary Data 8). This was because a larger number of samples in the placental mQTL dataset is likely to result in a greater amount and more significant set of mQTLs for SMR. Most neurological disorders, except for epilepsy, showed a higher proportion of significant SMR hits in placenta compared to blood. For instance, Parkinson’s and Autism spectrum disorder (ASD), together with birthweight (BW), represented the traits with the highest ratios within these categories, respectively. In contrast, most immune traits, except for Hashimoto’s thyroiditis showed a higher proportion of SMR hits in blood (< 1 enrichment ratios), as we would expect considering the biology of these traits.

To provide insights into the functional relevance of the placental SMR hits, we examined expression quantitative trait methylation sites (eQTMs) from a set of 195 fetal placenta samples from the RICHS study [20]. Body measurements were the traits with the largest numbers of SMR hits that were also placental eQTMs (Fig. 2C, Supplementary Data 8, 10). Yet, when considering the proportion of SMR hits that are placental eQTMs, neurological and digestive disorders were more prominent (Fig. 2D).

Lastly, we performed heritability enrichment analyses with LDAK [21] using placental and whole blood mQTL annotations [19] (Supplementary Data 11). Whole blood showed higher absolute enrichment estimates than placenta, particularly for immune-related traits (Fig. 3A). Notably, the placental mQTL resource included substantially more SNP-CpG associations than whole blood (44.5 million versus 19.3 million associations), despite only a modest difference in the number of unique SNPs (4.24 versus 3.49 million). Since the LDAK framework models annotations at the SNP level, without explicitly accounting for the number of CpG sites associated with each variant, these enrichment estimates may not fully capture differences in the regulatory connectivity of the underlying mQTL architectures across tissues. Correspondingly, differences between tissues were attenuated when considering enrichment Z-scores, which reflect the statistical robustness of the enrichment estimates (Fig. 3B). Placental mQTLs showed robust enrichment Z-scores across several trait domains, particularly neurological and anthropometric traits. Collectively, these results support tissue-specific differences in the genetic architecture captured by placental and blood mQTL annotations. This may reflect a stronger heritability enrichment from blood mVariants, and a broader distribution of pleiotropic regulatory contributions through DNAm in the placenta.

**Figure 3.**
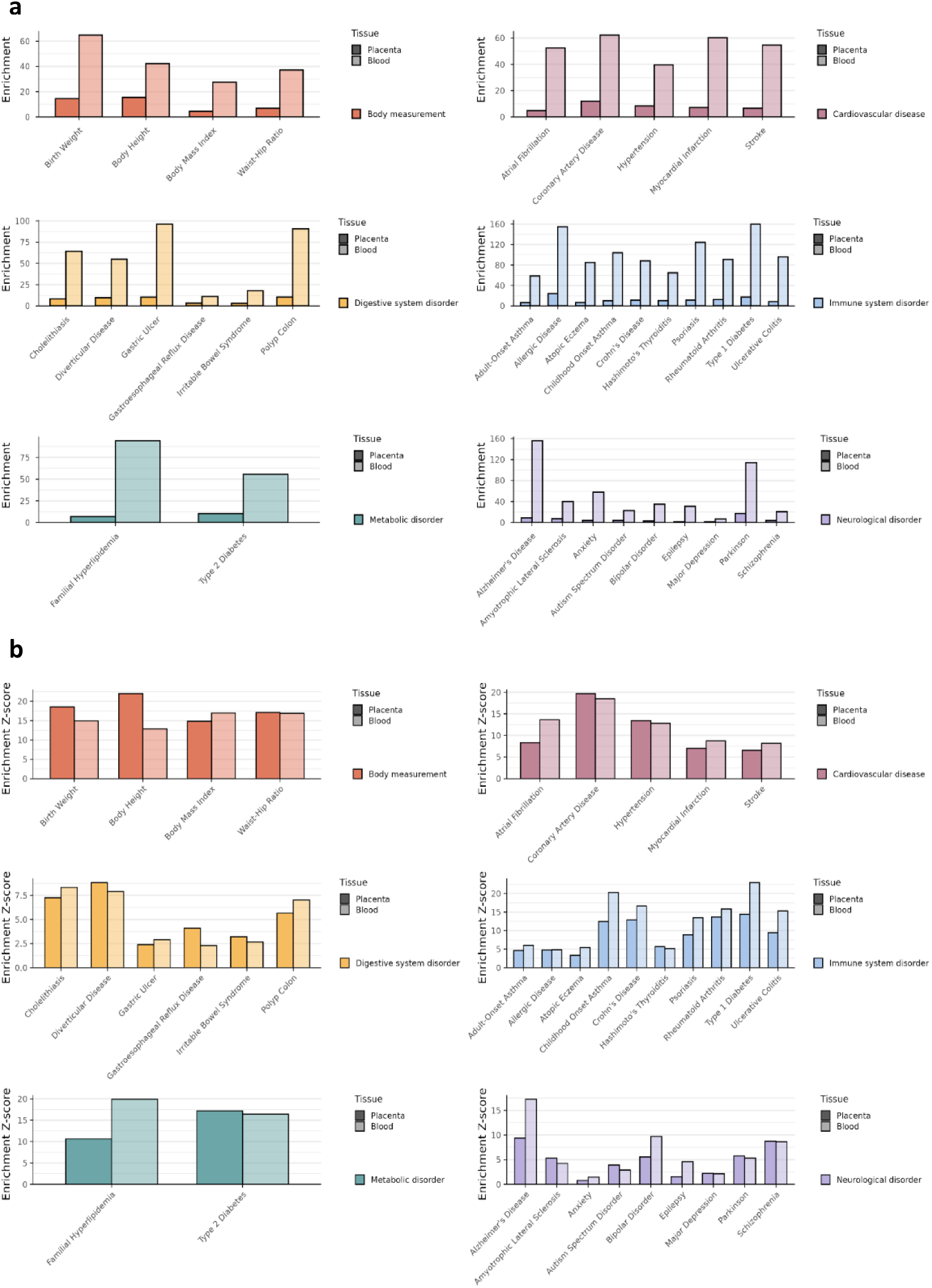
Heritability enrichment results from the 36 complex traits. Each panel groups the traits from a specific parent term, including body measurements, digestive system disorders, metabolic disorders, cardiovascular diseases, immune system disorders and neurological disorders. a) Shows the heritability enrichment across complex disease conditions while b) shows the Z-score of the enrichment. In all the plots X-axes represent the individual traits, and the darker bar represents placenta while the lighter one shows values calculated in whole blood.

Body measurements constitute the parent term with the largest number of placental SMR hits, as well as the highest absolute number overlapping placental eQTMs. Yet, these were less prominent after considering SMR enrichment ratios and the proportion of hits supported by eQTMs. In turn, neurological disorders show enrichment ratios consistently above one, indicating a relative overrepresentation of trait-associated signals in placenta compared with whole blood. Within this group, SCZ presents the largest number of SMR hits and placental eQTMs, whereas ASD and Parkinson’s disease display the highest enrichment ratios, and amyotrophic lateral sclerosis (ALS) shows the highest proportion of SMR hits supported by placental eQTMs. As expected, blood DNAm appears more informative for immune-related traits, with the notable exception of Hashimoto’s thyroiditis. Together, these patterns suggest that placental DNAm captures distinct architectures of genetic susceptibility across body systems, rather than acting as a uniform mediator.

Birth weight (BW) shows the highest SMR enrichment ratio in placenta among body measurements, together with a relatively high enrichment of the explained heritability in this organ. BW is strongly influenced by the intrauterine environment and is a well-established predictor of later-life health [22]. Using placental eQTM data, we identified *IP6K3* as a novel candidate gene regulated by 8 different CpG sites in placenta that were pleiotropically associated with BW, body height, BMI, type 1 and 2 diabetes, and coronary artery disease. Fig. 4 depicts a 300 kb genomic segment of regional pleiotropy, with *IP6K3* placental expression-associated CpGs operating under a shared genetic basis that confers susceptibility to anthropometric and cardiometabolic traits. *IP6K3* belongs to the inositol hexakisphosphate kinase family, involved in cellular signaling and energy metabolism [23]. In mouse models, *IP6K3* has been proposed as a target in obesity and metabolic disease [24, 25] and is a paralogue of *IP6K1*, a known regulator of insulin signaling [26]. We also identified cg08724371, correlated to the expression levels in placenta of the adjacent gene *FES*, also a placental eQTL. *FES* has previously been proposed as a functional effector gene for BW through placental DNAm and transcriptomic regulation [15]. Notably, cg08724371 was also pleiotropically associated with myocardial infarction for the first time in the present study, consistent with epidemiological evidence linking fetal growth to cardiometabolic disease risk later in life [27, 28]. These observations are consistent with a shared developmental architecture linking fetal growth and cardiometabolic disease susceptibility, through novel and known mQTL linked genes, some with translational potential.

**Figure 4.**
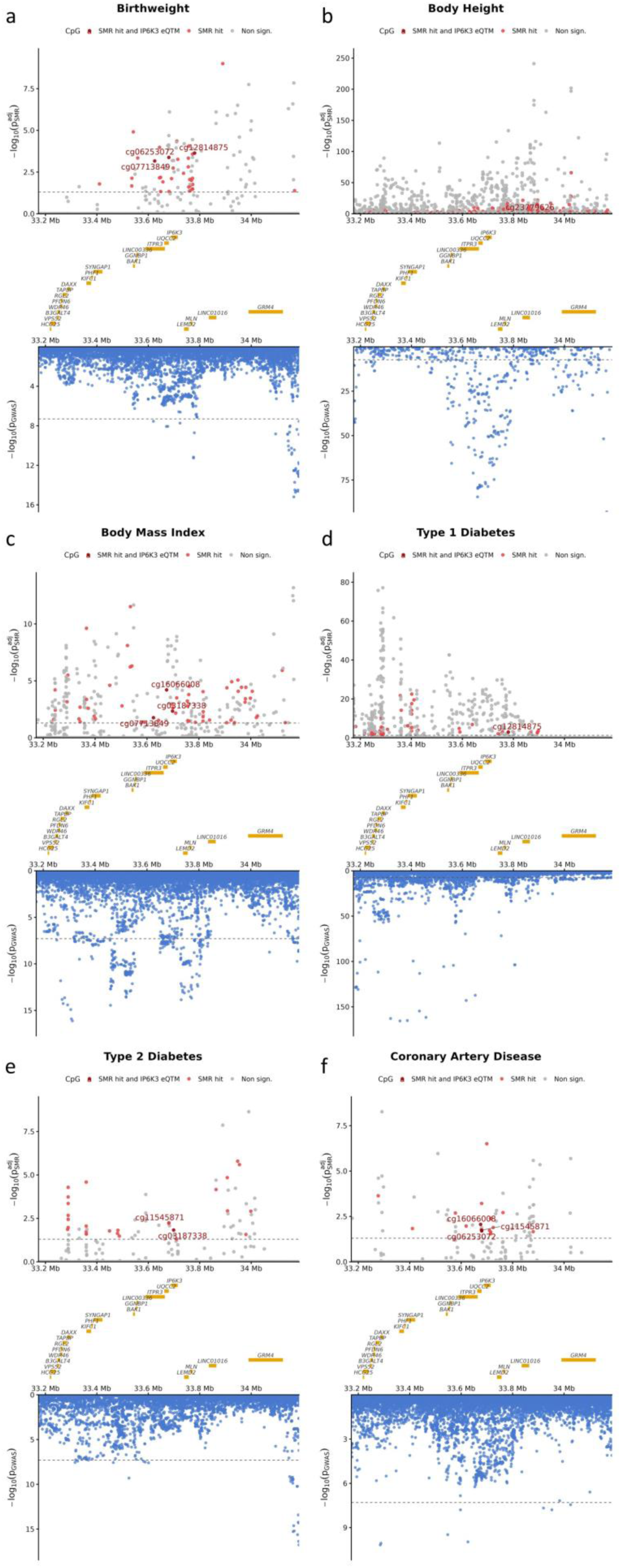
SMR and GWAS results for the genomic region containing *IP6K3*. Each panel shows a trait for which SMR-significant CpGs are associated with placental *IP6K3* expression: a) BW, b) body height, c) BMI, d) type 1 diabetes, e) type 2 diabetes and f) coronary artery disease. In all panels, the top plot displays SMR results, where the Y-axis represents –log10 Bonferroni P_SMR_ values and the X-axis shows the genomic coordinates. The dashed horizontal line indicates the significance threshold (Bonferroni P_SMR_ = 0.05). Each dot represents a CpG, colored according to HEIDI and SMR test results: grey indicates CpGs failing the HEIDI and/or SMR test, light red indicates CpGs passing both the HEIDI and SMR test, and dark red indicates CpGs passing both tests and identified as *IP6K3* eQTMs. The bottom plot shows the corresponding GWAS results for the region, where the Y-axis represents –log10(GWAS P-value) and the X-axis shows chromosome 6 genomic coordinates. The dashed line marks the genome-wide significance threshold (P = 5 × 10⁻⁸). The region between the two plots illustrates the gene distribution across the genomic interval.

Given its central role in fetal development, the placenta is an important contributor to neurodevelopment [29]. We identified regional pleiotropy in a 0.5 Mb genomic segment overlapping the gene promoter and body of *RGS6*. Placental DNAm at 28 CpGs in this region were pleiotropically associated with SCZ and correlated with *RGS6* placental expression, showing a potentially causal relationship between placenta DNAm and the disorder (Fig. 5). *RGS6* has previously been linked with central nervous system, anxiety, mood disorders and alcohol consumption [30], even as a potential therapeutic target [31], but not with placental biology. Beyond novel findings, we also replicated associations of *NAGA*, *VPS37B* and *TOB2P1* with SCZ, previously postulated as potentially causal in SCZ through DNAm-regulated placental gene expression [13], consistent with the robustness of the resource.

**Figure 5.**
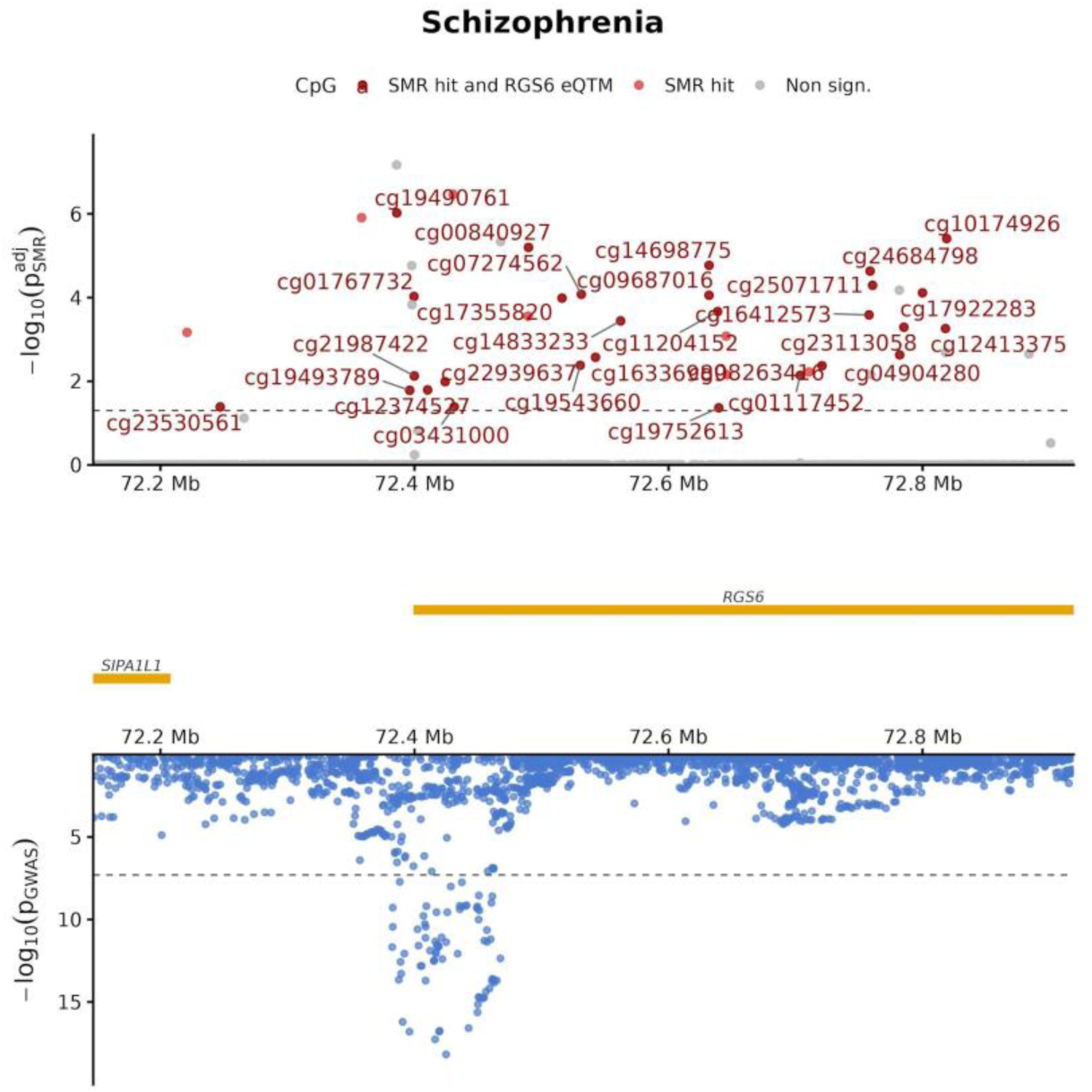
SMR and GWAS results for the genomic region containing *RGS6*. The top plot displays SCZ SMR results, where the Y-axis represents –log10 Bonferroni P_SMR_ values and the X-axis shows the chromosome 14 genomic coordinates. The dashed horizontal line indicates the significance threshold (Bonferroni P_SMR_ = 0.05). Each dot represents a CpG, colored according to HEIDI and SMR test results: grey indicates CpGs failing the HEIDI and/or SMR test, light red indicates CpGs passing both the HEIDI and SMR test, and dark red indicates CpGs passing both tests and identified as *RGS6* eQTMs. The bottom plot shows the corresponding GWAS results for the region, where the Y-axis represents –log10(GWAS P-value) and the X-axis shows chromosome 14 genomic coordinates. The dashed line marks the genome-wide significance threshold (P = 5 × 10⁻⁸). The region between the two plots illustrates the gene distribution across the genomic interval.

In addition, we found shared regulatory architecture across trait domains. Particularly in major depressive disorder (MDD), we identified cg10318063, associated with the disease in a previous placental study [13] and correlated with placental *LRFN5* expression according to our eQTM analysis. In the present study, *LRFN5* expression was associated with eight additional CpG sites pleiotropically linked to MDD. Four of these were also associated with gastroesophageal reflux disorder, suggesting coordinated regulation across neuropsychiatric and gastrointestinal traits. Such regional pleiotropy supports models of shared vulnerability rather than single-trait regulatory effects and is consistent with emerging biological frameworks linking gut and brain function [32].

Beyond trait-specific architecture, the distinctive epigenetic landscape of the placenta provides resolution for epigenetic mapping independently of assumptions about developmental timing. We identified a locus overlapping the first intron of *C9orf72*, comprising two placental CpGs pleiotropically associated with ALS. The most common genetic cause of familial ALS and frontotemporal dementia is a hexanucleotide repeat expansion in the first exon of *C9orf72*, often accompanied by promoter hypermethylation [33]. Although DNAm at ALS-associated CpGs was not correlated with placental *C9orf72* expression, this finding indicates that placental DNAm can capture disease-relevant regulatory signals independently of transcriptional coupling, highlighting the value of the placental DNAm-based resolution even when the etiologically relevant window cannot be assigned to prenatal life. In contrast, *AP3B2* emerges as an ALS candidate with functional support at the level of gene expression in the placenta, as six CpG sites were jointly associated with both the disease and *AP3B2* expression. *AP3B2* has previously been implicated in early-onset epileptic encephalopathy and autoimmune cerebellar ataxia [34, 35], but not in ALS. Together, these findings illustrate how integration of placental mQTL and eQTM signals can help distinguish disease-relevant DNAm variation from *loci* more plausibly linked to prenatal regulatory mechanisms.

Several limitations should be considered. The inclusion of cohorts profiled with the 450k array reduced power for sites unique to the EPIC array. eQTM analyses were restricted to a single cohort, potentially introducing cohort-specific effects and resulting in a reduced statistical power for this part of the pipeline. Moreover, SMR cannot distinguish between causal and pleiotropic associations, and results should be interpreted accordingly. Finally, sex-specific and cell-type specific mQTLs were not examined, despite evidence of sex-specific effects and placental cellular heterogeneity.

In summary, this placental *cis*-mQTL resource provides a high-resolution framework for studying the genetic regulation of DNAm in the context of early development. By leveraging the placenta’s distinctive epigenetic architecture, our analyses reveal trait-specific patterns of regulatory enrichment, highlight disease-relevant DNAm signals that are not necessarily coupled to placental gene expression, and identify regions of shared regulatory vulnerability across complex traits. Together, these results provide a structured basis for future integrative analyses aimed at refining the timing, tissue context, and mechanisms through which genetic and epigenetic variation contribute to complex disease susceptibility.

## Data Availability

All data produced are available online at https://pace-placenta-mqtl.streamlit.app/.

## Acknowledgements

We would like to thank all the participants and their families, as well as all PACE investigators, for their generous contribution.

## Author’s contributions

N.F.-J., C.L., M.B., J.R.B., and A.C.-P. conceived and designed the study. A.C.-P., I.G.-M., H.S., S.M., A.H.-L, L.B., M.G., D.C., J.T., C.B., J.D., and M.B. carried out data analysis. T.M.E., J.T., M.C.-T., M.L., K.H., J.T., M.D.F., R.J.S., J.F.D., S.A.-L., P.-E.J., S.H., A.I., M.L.-P., K.M.B., J.D., J.L., M.V., K.R., M.F.-H., J.S., B.H., J.L., S.J.L., J.C., M.B., C.J.M., J.R.B., C.L., and N.F.-J. actively participated in patient recruitment and/or data acquisition. S.J.L. was supported by the Intramural Research Program of the NIH, National Institute of Environmental Health Sciences. The contributions of the NIH author(s) are considered Works of the United States Government. The findings and conclusions presented in this paper are those of the author(s) and do not necessarily reflect the views of the NIH or the U.S. Department of Health and Human Services. A.C.-P., N.F.-J., and C.L. interpreted the results and wrote the first draft of the manuscript. N.F.-J. and C.L. jointly directed this project. All authors made substantial contributions to the acquisition, analysis, or interpretation of data, and read and critically revised the manuscript.

## Bibliography

1. Barker, D.J., The origins of the developmental origins theory. J Intern Med, 2007. 261(5): p. 412–7.

2. O’Brien, K. and Y. Wang, The Placenta: A Maternofetal Interface. (1545-4312 (Electronic)).

3. Cohen, N.J., et al., Placental DNA methylation key topics: sex– and cell-type specificity, mediation, multi-omics, and biomarker discovery. (1750-192X (Electronic)).

4. Lapehn, S. and A.G. Paquette, The Placental Epigenome as a Molecular Link Between Prenatal Exposures and Fetal Health Outcomes Through the DOHaD Hypothesis. Curr Environ Health Rep, 2022. 9(3): p. 490–501.

5. Diez-Ahijado, L., et al., Evaluating the association between placenta DNA methylation and cognitive functions in the offspring. Transl Psychiatry, 2024. 14(1): p. 383.

6. Patel, P., et al., Association between placental epigenetic age acceleration and early postnatal growth patterns. Sci Rep, 2025. 15(1): p. 29597.

7. Breton, E., et al., Placental NEGR1 DNA methylation is associated with BMI and neurodevelopment in preschool-age children. Epigenetics, 2020. 15(3): p. 323–335.

8. Everson, T.M., et al., Placental DNA methylation signatures of maternal smoking during pregnancy and potential impacts on fetal growth. Nat Commun, 2021. 12(1): p. 5095.

9. Fernandez-Jimenez, N., et al., A meta-analysis of pre-pregnancy maternal body mass index and placental DNA methylation identifies 27 CpG sites with implications for mother-child health. Commun Biol, 2022. 5(1): p. 1313.

10. Oliva, M., et al., DNA methylation QTL mapping across diverse human tissues provides molecular links between genetic variation and complex traits. Nat Genet, 2023. 55(1): p. 112–122.

11. Min, J.L., et al., Genomic and phenotypic insights from an atlas of genetic effects on DNA methylation. Nat Genet, 2021. 53(9): p. 1311–1321.

12. Casazza, W., et al., Sex-dependent placental methylation quantitative trait loci provide insight into the prenatal origins of childhood onset traits and conditions. iScience, 2024. 27(2): p. 109047.

13. Cilleros-Portet, A., et al., Potentially causal associations between placental DNA methylation and schizophrenia and other neuropsychiatric disorders. Nat Commun, 2025. 16(1): p. 2431.

14. Delahaye, F., et al., Correction: Genetic variants influence on the placenta regulatory landscape. PLoS Genet, 2019. 15(4): p. e1008118.

15. Tekola-Ayele, F., et al., Placental multi-omics integration identifies candidate functional genes for birthweight. Nat Commun, 2022. 13(1): p. 2384.

16. Do, C., et al., Mechanisms and Disease Associations of Haplotype-Dependent Allele-Specific DNA Methylation. Am J Hum Genet, 2016. 98(5): p. 934–955.

17. Bhattacharya, A., et al., Placental genomics mediates genetic associations with complex health traits and disease. Nat Commun, 2022. 13(1): p. 706.

18. Mägi, R. and A.P. Morris, GWAMA: software for genome-wide association meta-analysis. BMC Bioinformatics, 2010. 11: p. 288.

19. Hannon, E., et al., Leveraging DNA-Methylation Quantitative-Trait Loci to Characterize the Relationship between Methylomic Variation, Gene Expression, and Complex Traits. Am J Hum Genet, 2018. 103(5): p. 654–665.

20. Appleton, A.A., et al., Prenatal Programming of Infant Neurobehaviour in a Healthy Population. Paediatr Perinat Epidemiol, 2016. 30(4): p. 367–75.

21. Speed, D. and D.J. Balding, SumHer better estimates the SNP heritability of complex traits from summary statistics. Nat Genet, 2019. 51(2): p. 277–284.

22. Belbasis, L., et al., Birth weight in relation to health and disease in later life: an umbrella review of systematic reviews and meta-analyses. BMC Med, 2016. 14(1): p. 147.

23. Shears, S.A.-O., Intimate connections: Inositol pyrophosphates at the interface of metabolic regulation and cell signaling. (1097-4652 (Electronic)).

24. Mukherjee, S., J. Haubner, and A. Chakraborty, Targeting the Inositol Pyrophosphate Biosynthetic Enzymes in Metabolic Diseases. LID – 10.3390/molecules25061403 [doi] LID– 1403. (1420-3049 (Electronic)).

25. Moritoh, Y., et al., Inositol Hexakisphosphate Kinase 3 Regulates Metabolism and Lifespan in Mice. (2045-2322 (Electronic)).

26. Rajasekaran, S.S., et al., Inositol hexakisphosphate kinase 1 is a metabolic sensor in pancreatic β-cells. (1873-3913 (Electronic)).

27. Raisi-Estabragh, Z., et al., Lower birth weight is linked to poorer cardiovascular health in middle-aged population-based adults. Heart, 2023. 109(7): p. 535–541.

28. Barker, D.J., et al., Weight in infancy and death from ischaemic heart disease. The Lancet, 1989. 334(8663): p. 577–580.

29. Rosenfeld, C.S., The placenta-brain-axis. J Neurosci Res, 2021. 99(1): p. 271–283.

30. Stewart, A., et al., Regulator of G-protein signaling 6 (RGS6) promotes anxiety and depression by attenuating serotonin-mediated activation of the 5-HT(1A) receptor-adenylyl cyclase axis. (1530-6860 (Electronic)).

31. Ahlers, K.E., B. Chakravarti, and R.A.-O. Fisher, RGS6 as a Novel Therapeutic Target in CNS Diseases and Cancer. (1550-7416 (Electronic)).

32. Gong, W., et al., Role of the Gut-Brain Axis in the Shared Genetic Etiology Between Gastrointestinal Tract Diseases and Psychiatric Disorders: A Genome-Wide Pleiotropic Analysis. (2168-6238 (Electronic)).

33. Sellier, C., et al., C9ORF72 hexanucleotide repeat expansion: From ALS and FTD to a broader pathogenic role? (0035-3787 (Print)).

34. Assoum, M., et al., Autosomal-Recessive Mutations in AP3B2, Adaptor-Related Protein Complex 3 Beta 2 Subunit, Cause an Early-Onset Epileptic Encephalopathy with Optic Atrophy. (1537-6605 (Electronic)).

35. Honorat, J.A., et al., Autoimmune gait disturbance accompanying adaptor protein-3B2-IgG. (1526-632X (Electronic)).

