## Supplementary material for "Placental DNA methylation captures shared and trait-specific genetic susceptibility across complex health conditions": Methods

### Study cohorts

All the participating cohorts were involved in the Pregnancy And Childhood Epigenetics (PACE) consortium. The nine cohorts with genotype and DNAm data from placental tissue were Barcelona Life Study Cohort (BiSC) [1], Early Autism Risk Longitudinal Investigation (EARLI) [2], Study on the pre- and early postnatal determinants of child health and development (EDEN) [3], Genetics of Glucose Regulation in Gestation and Growth (Gen3G) [4], *Infancia y Medio Ambiente* or Environment and Childhood (INMA) project [5], Intrauterine Sampling in Early Pregnancy Study (ITU) [6], Markers of Autism Risk in Babies-Learning Early Signs (MARBLES) [7], Prediction and Prevention of Preeclampsia and Intrauterine Growth Restriction (PREDO) [8] and Rhode Island Child Health Study (RICHS) [9]. For instance, EDEN cohort was divided in two groups, EPIMEX and FDF, according to the DNAm array used (see *Preprocessing of DNAm data* section for more details). Subsequently, 10 studies were meta-analyzed, although they came from 9 different cohorts. For all cohorts, participants provided informed consent prior to data and sample collection, and the study protocols received approval from the respective Institutional Ethics Committees. Exclusion criteria for this study were non-singleton births, pre-eclampsia or several congenital malformations. Detailed methods for each cohort are provided in Supplementary Note and Supplementary Data 1.

### Preprocessing of genotype data

DNA extracted from placental or cord blood samples was used to generate fetal genotype data assessed with genotyping Illumina arrays or whole genome sequencing. See Supplementary Note for details on placenta and blood collection, DNA extraction and genotype acquisition in each cohort. Genotype data quality control was standardized across all cohorts and performed with PLINK 1.9 software, following the standard recommendations [10], [11], [12]. Variants were excluded if they exhibited a genotype call rate < 95%, a minor allele frequency (MAF) <1%, or deviation from Hardy–Weinberg equilibrium (HWE) with an exact test $P < 1 \times 10^{-6}$. Sample-level quality control removed individuals with discordant genetic and reported sex, mean heterozygosity values exceeding ±4 standard deviations from the cohort mean, or >3% missing genotypes. Identity-by-descent (IBD) was assessed using PLINK, and within pairs of individuals with PI-HAT > 0.18, the sample showing the greater proportion of missing genotypes was excluded.

Cohort's final dataset was processed with Will Rayner's preparation Perl script, as recommended by the Michigan Imputation Server developers [13]. Before imputation, data was converted into VCF format. Phasing of haplotypes was done with Eagle v2.4 [14] and genotype imputation with Minimac4 [15], both implemented in the code from the imputation server. Genotype files were imputed locally or remotely with the Michigan Imputation Server using the (HRC) [16] or the 1000 Genomes project (1000G) [17] as reference panels. Before imputation, data was converted into VCF format. Phasing of haplotypes was done with Eagle v2.4 [16] and genotype imputation with Minimac4 [17], both implemented in the code from the imputation server. Finally, we removed variants with an imputation $R^2 < 0.9$, a MAF lower than 5% and a HWE $P < 0.05$. Genotypes were reported based on the GRCh37 reference genome. Only those samples with paired placental DNAm data were considered in this analysis. More details on the final sample size and specific methods from each cohort can be found in Supplementary Note and Supplementary Data 1.

### Preprocessing of DNAm data

Placental DNAm was assessed with Illumina Infinium HumanMethylation450 or 450K array (EDEN - FDF, EARLI and RICHS, N=612) or with Illumina MethylationEPIC BeadChip or EPIC array (BiSC, EDEN -

EPIMEX, Gen3G, INMA, ITU, MARBLES and PREDO, N=1,971). See supplementary Note for details on DNAm data quality control and normalization in each cohort. Quality control of the DNAm data was standardized across all cohorts and performed with the PACEanalysis R package (v.0.1.7) [18], [19]. It included the following steps: discarding samples with call rate below 95%, with sex inconsistencies, duplicates and those with DNA contamination (from another subject or the mother). Only those samples with paired child genotype data were considered in this study. The quality control also excluded probes with a call rate lower than 95%, in the sex chromosomes, with SNPs (European, MAF $\geq$ 5%) and cross-hybridizing potential [20].

Study-specific datasets were normalized with Noob (normal-exponential out-of-band), a background correction method with dye-bias normalization, implemented in the minfi R package [21], [22], followed by the functional normalization method [23]. We corrected for type-2 probe bias values with the beta-mixture quantile (BMIQ) normalization [24]. Next, each cohort performed Principal Component Analysis (PCA) and tested the association of the 12 first PCs with the main biological and technical variables, including batch, sample's sex and race/ethnicity. Array batch effect was controlled with the ComBat method [25]. Extreme outliers were winsorized to the 1% percentile (0.5% on each side); percentiles were estimated with the empirical beta distribution. Lastly, we applied the rank-based inverse normal transformation (RNT) to methylation beta values before methylation quantitative trait methylation (mQTL) mapping.

To get placental cell type proportions, we used the placenta DNAm reference panel from the 3$^{rd}$ trimester implemented in the Planet R package [26]. Cell proportions for six populations were obtained: syncytiotrophoblasts (STB), trophoblasts (TB), nucleated red blood cells (RBCs), Hofbauer cells, endothelial cells, and stromal cells.

**Mapping of the placental mQTLs**

mQTL mapping was performed within each cohort separately using TensorQTL's nominal mode [27], which fits linear regressions between genotypes and normalized DNAm Rank-normal transformed (RNT) values. The *cis*-region was set at ±0.5Mb from each CpG site, consistent with previous studies [19], [28], [29], [30], [31]. The covariates included in the regression model were the sex of the fetus, the genotype PCs, the non-genetic DNAm PCs, and the cell type proportions. The number of genetic and DNAm PCs to be included as covariates in the model was determined independently within each cohort, depending on sample characteristics (details in Supplementary Note). Following GoDMC guidelines and previous work from our Group [19], [32], to prevent multicollinearity between DNAm PCs and other covariates, we calculated DNAm PCs on the residuals from a multiple linear regression of the DNAm RNT-values adjusted by covariates (sex of the fetus, 5 genotype PCs, and the six estimated cell types). Each cohort kept the number of DNAm PCs that cumulatively explained 80% of the variance with a maximum number of 20 PCs for subsequent steps. Then a genome-wide association study (GWAS) was performed for each of the DNAm PCs and only those PCs that were not associated with any SNP at a suggestive threshold ($P > 1 \times 10^{-7}$) were retained for mQTL mapping.

**Preprocessing of placental mQTL summary statistics across cohorts**

We performed quality control of the results from the cohorts (mQTL summary statistics) using the EasyQC2 R package [33], which harmonized summary statistics to ensure comparability across cohorts before meta-analysis. Amongst the quality control steps, we removed mQTLs with invalid or missing statistics, as well as associations involving monomorphic variants, or variants with missing or non-canonical alleles (i.e., alleles other than A, C, G, T or an INDEL). We removed variants with mismatching reference allele to the

HRC imputation panel. To confirm that all remaining genetic variants had a MAF > 5% and HWE $P > 0.05$, we filtered the data using these thresholds. Finally, for each of the studies, we compared the effect allele frequency against HRC reference and computed λ values.

**Meta-analysis of placental mQTL summary statistics**

We performed inverse variance-weighted fixed effects meta-analysis using GWAMA [34]. Due to computational constraints, the meta-analysis was conducted in ten chunks per chromosome; subsequently, all the results were merged into a single dataset. Additionally, a shadow meta-analysis was conducted to validate the results, showing consistent findings with the primary meta-analysis. The primary placental mQTL database included all associations reported in more than one cohort and those probes with at least one *cis*-mQTL at $P < 5 \times 10^{-8}$. From this primary set, we also derived a secondary, more stringent database including only the top or lead mVariant per mSite (i.e., the variant with the lowest *P* per mSite). The *P*-value used in this analysis corresponds to the nominal *P*-value from the meta-analysis.

**Profiling of the meta-analyzed placental mQTLs**

We started by plotting distributions of the distance between mSite and mVariant pairs, the count of mQTLs detected across the cohorts, and the heterogeneity $I^2$ statistic. Next, we leverage R annotation packages IlluminaHumanMethylation450kanno.ilmn12.hg19 [35] and IlluminaHumanMethylationEPICanno.ilm10b4.hg19 [36], performed $\chi^2$-square tests to assess for enrichment or depletion of UCSC RefGene and Relation to CpG Island annotations. We used eFORGE v2.0 [37], [38], [39] to assess enrichment and depletion of overlap with tissue-specific regulatory features including chromatin all 15-state marks, DNase I hypersensitivity sites (DHS) and H3K4me1 histone marks from the consolidated ROADMAP Epigenomics Mapping Consortium [40]. Enrichment and depletion analyses for each of the three putative elements were conducted independently and benchmarked against the corresponding annotations from the consolidated ROADMAP Epigenomics reference panel. Following the recommendations of the eFORGE developers, we restricted analyses to the top 10,000 probes in order to prevent saturation of background bins, particularly of those linked to CpG island annotation categories.

We used the MissMethyl R package [41] to perform over-representation analyses leveraging Gene Ontology (GO) and Kyoto Encyclopedia of Genes and Genomes (KEGG) annotations. MissMethyl performs a hypergeometric test taking into account the bias derived from the differing number of probes per gene and the multiple genes annotated per CpG. ENCODE Transcriptional Factor Binding Sites (TFBS) enrichment analyses was conducted with the LOLA R package [42] with a Fisher's exact test considering the genes annotated in the Illumina annotation R packages.

**Genome-wide association studies**

Using the NHGRI-EBI GWAS Catalog v.1.0.2 [43], we selected complex traits with at least one GWAS with more than 50,000 samples, a balance between the sample size of cases and controls when applicable, imputed genotype, and full-summary statistics publicly available. None of our cohorts participated in the GWAS selected. We harmonized alleles to the HRC reference panel for the GWAS from each of the 36 complex traits selected, in order to make them comparable to the placental mQTL database (Supplementary Data 6). Finally, the summary statistics files were formatted for Summary-based Mendelian Randomization (SMR) and LDAK analyses.

**Multi-SNP-based Mendelian Randomization analysis**

We performed multi-SNP-based MR (SMR-multi) [44] analysis with *cis*-mQTL mVariants as instrumental variables (IVs), CpG methylation as the exposure (X), and complex traits as the outcome (Y), with the SMR software (v.1.3.1) [45]. SMR integrates GWAS and QTL summary statistics to test for pleiotropic associations between molecular quantitative traits (i.e. DNAm), and a complex phenotype. SMR-multi incorporates multiple variants at a *cis*-mQTL locus in the SMR-test to compute the causative effect of an exposure on an outcome ($b_{xy}$). Initially, SMR-multi selects all the variants with a meta-analysis $P < 5 \times 10^{-8}$ in the *cis* region (±0.5Mb of the CpG). Then, variants in linkage disequilibrium (LD) with the top associated SNP (LD $r^2 > 0.1$) are excluded to reduce potential multicollinearity. Next, causative effects ($b_{xy}$) from exposure on the outcome estimated for each SNP, are combined using an approximate set-based test accounting for LD among SNPs [44]. Finally, we performed the HEIDI test that uses multiple SNPs in a *cis*-mQTL region to distinguish pleiotropy from linkage.

This analysis was conducted separately on the placental mQTLs and a public whole blood mQTLs database from Hannon *et al.* 2018 [46]. We choose this study because it is the largest publicly available mQTL database on whole blood including all the probes from the Illumina EPIC array. In both analyses, significant pleiotropic associations between DNAm and the 36 complex traits were selected as those with Bonferroni-corrected $P_{SMR} < 0.05$ and $P_{HEIDI} > 0.05$ (not showing heterogeneity). Note that the major histocompatibility complex region from chromosome 6 has been excluded from this part of the analysis following developer's guidelines [45] to avoid misleading results coming from the high LD in that region.

**Heritability enrichment analysis**

The heritability enrichment analysis was conducted with SumHer within LDAK (v.6) [47]. We obtained lists of unique mVariants for the placenta and blood mQTLs databases [46] with a meta-analysis $P < 5 \times 10^{-8}$, and computed the taggings, which record the expected heritability tagged by each predictor. In this case, we considered a binary annotation with each SNP tagged indicating mQTL status in the corresponding tissue. We computed the enrichment of heritability in the 36 complex traits using the default options. Since we had overlapping SNPs between the two databases (blood and placenta), we used *"--annotation-number"* and *"--annotation-prefix"* options, following developer's guidelines. Lastly, we computed the Z-score from the enrichment $h^2_{SNP}$ value to account for the robustness of the enrichment calculation of each trait. Note that the major histocompatibility complex region from chromosome 6 has been excluded from this part of the analysis to avoid misleading results coming from the high LD in that region.

**RICHS eQTMs**

Expression quantitative trait methylation (eQTM) analysis was performed in a subset of RICHS participants (see Supplementary Note). Short-read placenta RNA-seq data from a subset of RICHS samples were obtained using Illumina Hi-Seq 2500 platform. Placental DNAm and RNA-seq data from 195 samples were used to compute eQTMs using linear models in MatrixEQTL, considering a *cis* window of 0.5 Mb up and downstream of each CpG. Linear regressions were adjusted for sex, five PCs of gene expression, and the placenta cell type proportions estimated from DNAm data (planet R package). Results were corrected with false discovery rate (FDR).

**Data availability**

The summary statistics from the meta-analyses, the primary and secondary mQTL databases, as well as, all the associations regardless of statistical significance are publicly available: https://pace-placenta-mqtl.streamlit.app/.

Individual genotype and DNAm raw data are not publicly available because neither children nor their parents have given consent for open publication of individual-level and sensitive data. However, these data are accessible upon request, through external collaboration with the individual cohorts.

### Code availability

The code for the genotype and methylation QC, as well as the TensorQTL nominal mapping, is available in this GitHub repository link: https://github.com/ariadnacilleros/Cis-mQTL-mapping-protocol-formethylome, and it is also available in Zenodo [https://doi.org/10.5281/zenodo.14198427].

### References


1. Dadvand, P., et al., *Cohort Profile: Barcelona Life Study Cohort (BiSC).* Int J Epidemiol, 2024. **53**(3).
2. Newschaffer, C., et al., *P-068: The EARLI Study as a Resource for Research on Autism and the Environment.* Epidemiology, 2012. **23**(5S).
3. Heude, B., et al., *Cohort Profile: The EDEN mother-child cohort on the prenatal and early postnatal determinants of child health and development.* Int J Epidemiol, 2016. **45**(2): p. 353–63.
4. Guillemette, L., et al., *Genetics of Glucose regulation in Gestation and Growth (Gen3G): a prospective prebirth cohort of mother-child pairs in Sherbrooke, Canada.* BMJ Open, 2016. **6**(2): p. e010031.
5. Guxens, M., et al., *Cohort Profile: the INMA--INfancia y Medio Ambiente--(Environment and Childhood) Project.* Int J Epidemiol, 2012. **41**(4): p. 930–40.
6. Kvist, T., et al., *Cohort profile: InTraUterine sampling in early pregnancy (ITU), a prospective pregnancy cohort study in Finland: study design and baseline characteristics.* BMJ Open, 2022. **12**(1): p. e049231.
7. Hertz-Picciotto, I., et al., *A Prospective Study of Environmental Exposures and Early Biomarkers in Autism Spectrum Disorder: Design, Protocols, and Preliminary Data from the MARBLES Study.* Environmental Health Perspectives, 2018. **126**(11): p. 117004.
8. Girchenko, P., et al., *Cohort Profile: Prediction and prevention of preeclampsia and intrauterine growth restriction (PREDO) study.* Int J Epidemiol, 2017. **46**(5): p. 1380–1381g.
9. Appleton, A.A., et al., *Prenatal Programming of Infant Neurobehaviour in a Healthy Population.* Paediatr Perinat Epidemiol, 2016. **30**(4): p. 367–75.
10. Anderson, C.A., et al., *Data quality control in genetic case-control association studies.* Nat Protoc, 2010. **5**(9): p. 1564–73.
11. Purcell, S., et al., *PLINK: a tool set for whole-genome association and population-based linkage analyses.* Am J Hum Genet, 2007. **81**(3): p. 559–75.
12. Chang, C.C., et al., *Second-generation PLINK: rising to the challenge of larger and richer datasets.* Gigascience, 2015. **4**: p. 7.
13. Das, S., et al., *Next-generation genotype imputation service and methods.* Nat Genet, 2016. **48**(10): p. 1284–1287.
14. Loh, P.R., et al., *Reference-based phasing using the Haplotype Reference Consortium panel.* Nat Genet, 2016. **48**(11): p. 1443–1448.
15. Fuchsberger, C., G.R. Abecasis, and D.A. Hinds, *minimac2: faster genotype imputation.* Bioinformatics, 2015. **31**(5): p. 782–4.
16. McCarthy, S., et al., *A reference panel of 64,976 haplotypes for genotype imputation.* Nat Genet, 2016. **48**(10): p. 1279–83.

17. Auton, A., et al., *A global reference for human genetic variation.* Nature, 2015. **526**(7571): p. 68–74.
18. Binder, A.M. *PACE Analyses*. Available from: https://www.epicenteredresearch.com/pace/.
19. Cilleros-Portet, A., et al., *Potentially causal associations between placental DNA methylation and schizophrenia and other neuropsychiatric disorders.* Nat Commun, 2025. **16**(1): p. 2431.
20. McCartney, D.L., et al., *Identification of polymorphic and off-target probe binding sites on the Illumina Infinium MethylationEPIC BeadChip.* Genom Data, 2016. **9**: p. 22–4.
21. Fortin, J.P., T.J. Triche, Jr., and K.D. Hansen, *Preprocessing, normalization and integration of the Illumina HumanMethylationEPIC array with minfi.* Bioinformatics, 2017. **33**(4): p. 558–560.
22. Triche, T.J., Jr., et al., *Low-level processing of Illumina Infinium DNA Methylation BeadArrays.* Nucleic Acids Res, 2013. **41**(7): p. e90.
23. Fortin, J.P., et al., *Functional normalization of 450k methylation array data improves replication in large cancer studies.* Genome Biol, 2014. **15**(12): p. 503.
24. Teschendorff, A.E., et al., *A beta-mixture quantile normalization method for correcting probe design bias in Illumina Infinium 450 k DNA methylation data.* Bioinformatics, 2013. **29**(2): p. 189–96.
25. Johnson, W.E., C. Li, and A. Rabinovic, *Adjusting batch effects in microarray expression data using empirical Bayes methods.* Biostatistics, 2007. **8**(1): p. 118–27.
26. Yuan, V., et al., *Cell-specific characterization of the placental methylome.* BMC Genomics, 2021. **22**(1): p. 6.
27. Taylor-Weiner, A., et al., *Scaling computational genomics to millions of individuals with GPUs.* Genome Biol, 2019. **20**(1): p. 228.
28. Hannon, E., et al., *An integrated genetic-epigenetic analysis of schizophrenia: evidence for co-localization of genetic associations and differential DNA methylation.* Genome Biol, 2016. **17**(1): p. 176.
29. Gibbs, J.R., et al., *Abundant quantitative trait loci exist for DNA methylation and gene expression in human brain.* PLoS Genet, 2010. **6**(5): p. e1000952.
30. Drong, A.W., et al., *The presence of methylation quantitative trait loci indicates a direct genetic influence on the level of DNA methylation in adipose tissue.* PLoS One, 2013. **8**(2): p. e55923.
31. Olsson, A.H., et al., *Genome-wide associations between genetic and epigenetic variation influence mRNA expression and insulin secretion in human pancreatic islets.* PLoS Genet, 2014. **10**(11): p. e1004735.
32. Min, J.L., et al., *Genomic and phenotypic insights from an atlas of genetic effects on DNA methylation.* Nat Genet, 2021. **53**(9): p. 1311–1321.
33. Winkler, T.W., et al., *Quality control and conduct of genome-wide association meta-analyses.* Nat Protoc, 2014. **9**(5): p. 1192–212.
34. Mägi, R. and A.P. Morris, *GWAMA: software for genome-wide association meta-analysis.* BMC Bioinformatics, 2010. **11**: p. 288.
35. Hansen, K.D., *Annotation for Illumina's 450k methylation arrays*. 2025.
36. Hansen, K.D., *Annotation for Illumina's EPIC methylation arrays*. 2025.
37. Breeze, C.E., et al., *eFORGE: A Tool for Identifying Cell Type-Specific Signal in Epigenomic Data.* Cell Rep, 2016. **17**(8): p. 2137–2150.
38. Breeze, C.E., et al., *eFORGE v2.0: updated analysis of cell type-specific signal in epigenomic data.* Bioinformatics, 2019. **35**(22): p. 4767–4769.
39. Breeze, C.E., *Cell Type-Specific Signal Analysis in Epigenome-Wide Association Studies.* Methods Mol Biol, 2022. **2432**: p. 57–71.
40. Kundaje, A., et al., *Integrative analysis of 111 reference human epigenomes.* Nature, 2015. **518**(7539): p. 317–30.

41. Phipson, B., J. Maksimovic, and A. Oshlack, *missMethyl: an R package for analyzing data from Illumina's HumanMethylation450 platform.* Bioinformatics, 2016. **32**(2): p. 286–8.
42. Sheffield, N.C. and C. Bock, *LOLA: enrichment analysis for genomic region sets and regulatory elements in R and Bioconductor.* Bioinformatics, 2016. **32**(4): p. 587–9.
43. Buniello, A., et al., *The NHGRI-EBI GWAS Catalog of published genome-wide association studies, targeted arrays and summary statistics 2019.* Nucleic Acids Res, 2019. **47**(D1): p. D1005–d1012.
44. Wu, Y., et al., *Integrative analysis of omics summary data reveals putative mechanisms underlying complex traits.* Nat Commun, 2018. **9**(1): p. 918.
45. Zhu, Z., et al., *Integration of summary data from GWAS and eQTL studies predicts complex trait gene targets.* Nat Genet, 2016. **48**(5): p. 481–7.
46. Hannon, E., et al., *Pleiotropic Effects of Trait-Associated Genetic Variation on DNA Methylation: Utility for Refining GWAS Loci.* Am J Hum Genet, 2017. **100**(6): p. 954–959.
47. Speed, D. and D.J. Balding, *SumHer better estimates the SNP heritability of complex traits from summary statistics.* Nat Genet, 2019. **51**(2): p. 277–284.
