## Supplementary Figures for "Placental DNA methylation captures shared and trait-specific genetic susceptibility across complex health conditions"

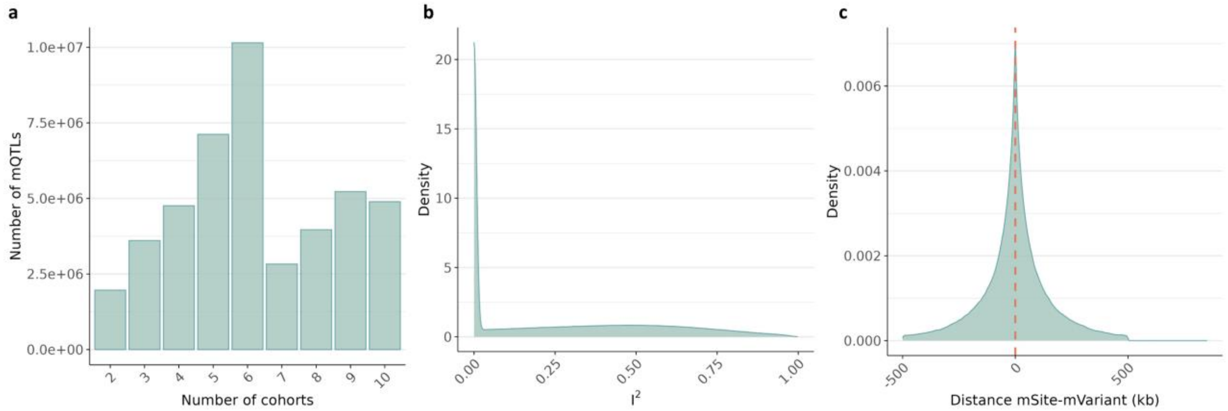


**Supplementary Figure 1. Quality control plots from GWAMA's results**. a) Barplot showing the number of placental mQTLs (Y-axis) from the primary database by the number of cohorts in which they were detected (X-axis). b) Density plot of the $I^2$ heterogeneity values of the primary database mQTLs. c) Density plot shows the distance between the mSite and mVariant in kb, represented on the X-axis. The red line corresponds to the mean of these values.

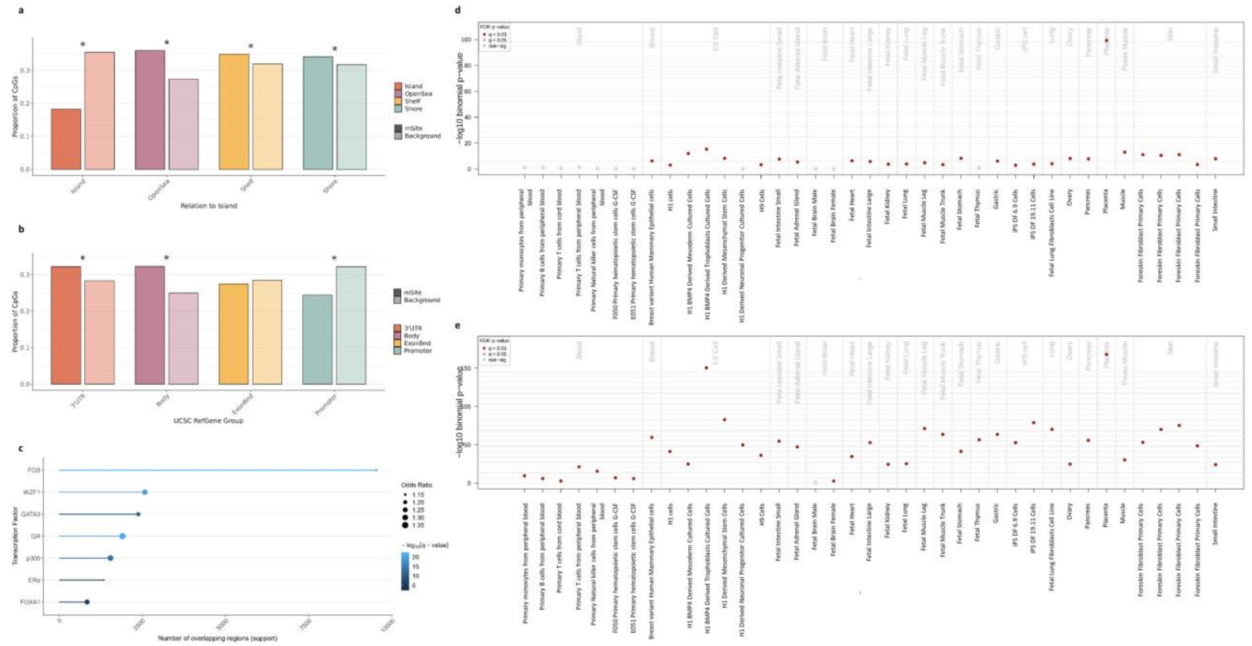


**Supplementary Figure 2. Characterization of the placental mQTLs.** Panel a) and b) show the proportion of mSites and background CpGs being defined by the Relation To Island and UCSC RefGene Group annotations from the Illumina annotation R packages. The "*" highlights significant enrichments or depletions regarding the Chi-square tests. Subfigure c) represents the results from the TFBS enrichment analysis performed with the LOLA R package. The eFORGE enrichment of DNase I hotspots and H3K4me1 broadPeaks considering the top 10,000 mSites are shown in panels d) and e), respectively. The Y-axis represents the −log10 P-value of the binomial enrichment test as suggested by the eFORGE developers, and the X-axis the tissues studied. FDR-corrected q-values below 0.01 and 0.05 are represented by red and pink dots, respectively, while blue dots show q-values > 0.05.

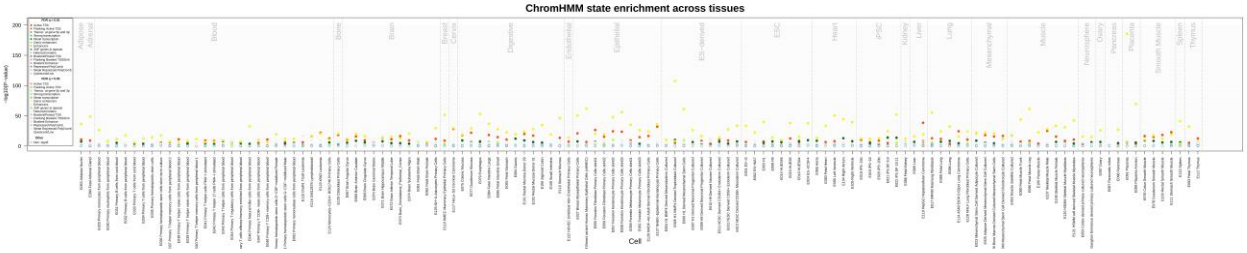


**Supplementary Figure 3. eFORGE enrichment of histone marks on the placental mQTLs.** The eFORGE enrichment of all histone marks broadPeaks considering the top 10,000 mSites is shown. The Y-axis represents the −log10 P-value of the binomial enrichment test as suggested by the eFORGE developers, and the X-axis the tissues studied. FDR-corrected q-values below 0.01 and 0.05 are represented by full and empty dots, respectively, color-coded according to the histone mark. While blue dots show q-values > 0.05.

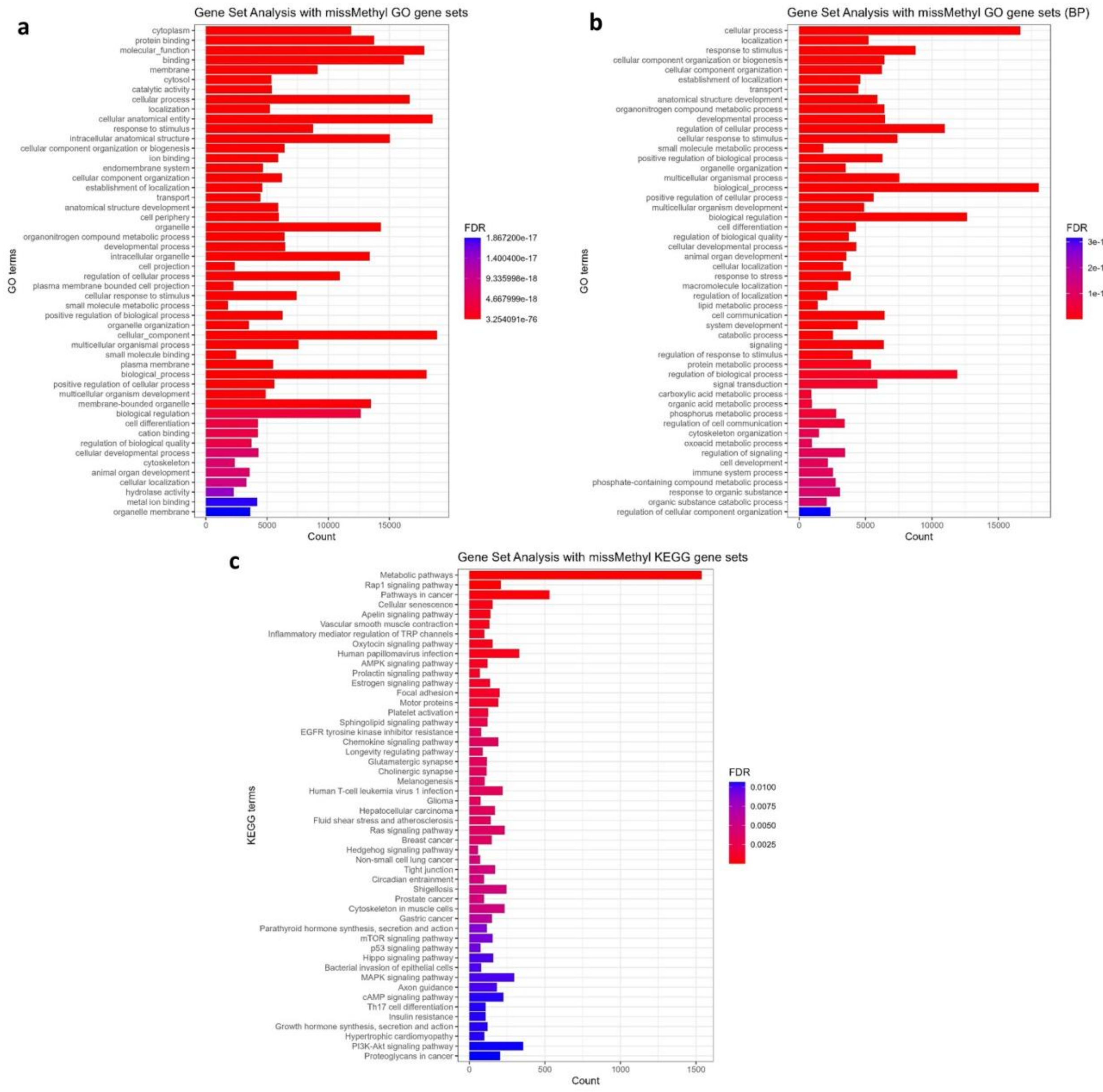


**Supplementary Figure 4. Over-representation and gene set enrichment analysis of the mSites.** An overrepresentation analysis was performed using MissMethyl R package and Gene Ontology (GO) and Kyoto Encyclopedia of Genes and Genomes (KEGG) gene sets. Results are plotted in the a, b and c barplots, respectively. In all three plots, the Y-axis represents the top gene sets enriched, and the adjusted p-value is color coded. The X-axes represent the counts of enriched genes.

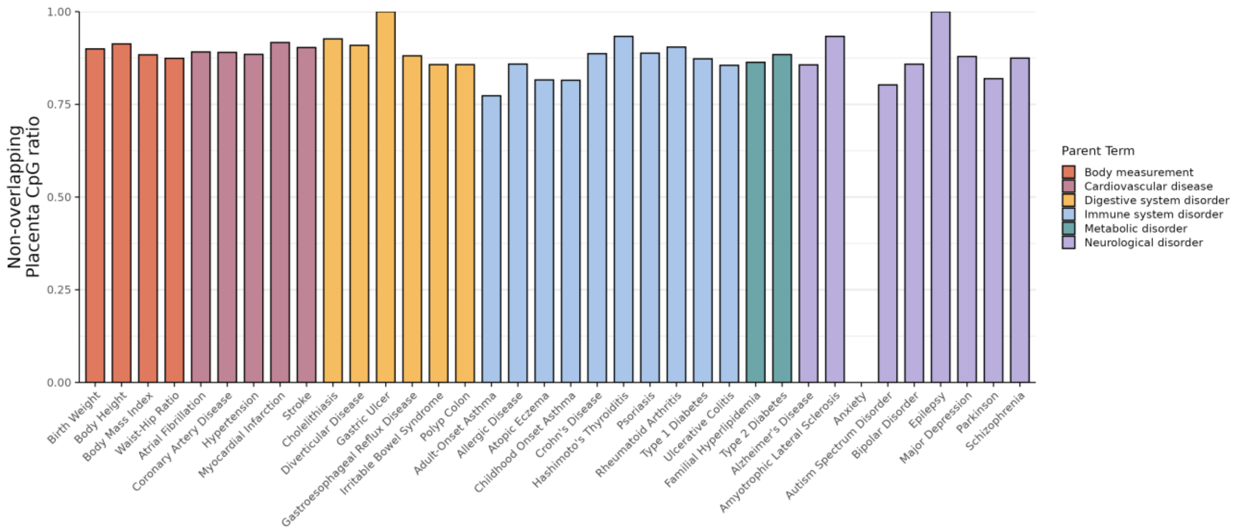


**Supplementary Figure 5. Proportion of overlapping placental SMR hits.** On the X-axis the 36 complex traits analyzed are shown and on the Y-axis the proportion from 0 to 1 of placenta SMR hits that are uniquely detected in this tissues compared to whole blood. Additionally, the complex traits have been color-coded as a function of their parent term.
