## Supplementary Note for "Placental DNA methylation captures shared and trait-specific genetic susceptibility across complex health conditions"

### Barcelona Life Study Cohort (BiSC)

#### Placental biopsies and DNA extraction

The Barcelona Life Study Cohort (BiSC) is a prospective cohort study of 1,080 pregnant women, their offspring and partners in Barcelona that aims to identify early environmental and genetic causes of normal and abnormal growth, development and health from foetal life until young adulthood [1]. Briefly, the enrolment of the BiSC participants was carried out between October 2018 and April 2021 at three tertiary university hospitals in Barcelona, Spain. Participants were recruited and had their first data collected at the end of the first trimester of pregnancy, with ups in the second and third trimesters of pregnancy, delivery, and months 1, 2, 6, 8, 12, 107 18, 24, and 48 postnatally. Pregnant women aged 18-45 with singleton pregnancies were included. Exclusions applied to those residing outside the catchment area, not being able to communicate effectively in Spanish, Catalan or English, or with fetuses having known congenital anomalies. The study was approved by the ethical committees of the centers involved in the study, and written informed consent was obtained from all the participants. The total number of mother-child pairs in BiSC was 1,032.

Out of the 1,032 mother-child pairs that were followed until birth, 611 placentas were collected based on maternal consent and the feasibility of collection at the hospital. Biopsies were obtained by trained gynaecologists following a harmonized protocol across hospitals. Briefly, placenta biopsies of around 2.5 cm (from the maternal to the foetal side) and 1 cm width were obtained from two opposite quadrants at a distance of around 3-4 cm from site of cord insertion. Then, these biopsies were cut in two, giving a total of 4 biopsies of 2.5 × 0.5 cm. Two of them (one from each quadrant) were directly frozen in liquid nitrogen and transferred to -80ºC. The other two biopsies were treated with RNAlater and sent to the laboratory where they were divided in four pieces of 0.5 × 0.5 cm, corresponding roughly to the foetal membranes, the upper chorionic villi, the lower chorionic villi and the maternal decidua. Finally, all the biopsies were stored at -80ºC for future use. Before DNA extraction, placenta samples were completely randomized and the distribution of main design and biological variables was checked across batches. For genomic DNA extraction, a fragment of approximately 5-6 cm3 (30-40 mg) was dissected from the chorionic villi biopsy below the foetal membranes collected in RNAlater. All the dissection process was done in liquid nitrogen to avoid the tissue thawed. Then, the tissue was disrupted/homogenized using a bead mill (bead beater) at 4ºC for 26 seconds. Genomic DNA was then isolated using the AllPrep®DNA/RNA/miRNA Universal Kit, (Qiagen, CA, USA). DNA was eluted in 80 µl and stored in different aliquots at -80ºC. DNA quality was evaluated on a NanoDrop spectrophotometer (Thermo Scientific, Waltham, MA, USA) and additionally 500 ng of DNA was run on 1% agarose gels to confirm that samples did not present visual signs of degradation.

Maternal (at 12 or 32 weeks of pregnancy or at delivery) and umbilical cord blood samples were collected in EDTA tubes (Vacutainer, Ref BD-368381). In the laboratory, EDTA tubes were centrifuged at 2000 g for 10 min and plasma, buffy coat and red cells were separated. DNA was extracted from 200 ul of buffy coat using the QIAsymphony platform and the QIAsymphony DSP DNA Mini Kit (Qiagen, Ref 937236) at the Hospital del Mar Research Institute (IMIM). DNA extraction was done separately for mothers and children, and, in both cases, samples were completely randomized.

#### Genotype data

A total of 1539 DNA samples (1513 unique) were selected for genotyping at the Spanish National Genotyping Centre (CEGEN) (Spain), including 589 maternal samples (976 unique) and 554 child samples (537 unique). Among the child DNA samples, la majority were obtained from umbilical cord blood and the rest from placenta. Maternal DNA samples were obtained from peripheral blood at any time point mentioned above. Genome-wide genotyping was conducted using the Infinium Global Screening (GSA) array v.3.0 with Multi-disease (MD) add-on content from Illumina, that contains 730,059 genetic variants (https://www.illumina.com/products/by-type/microarraykits/infinium-global-screening.html). Before genotyping DNA samples were quantified with the ADNds Quant-iT™ PicoGreen™ kit and an aliquot of 250 ng was transferred into the genotyping plate. Samples were genotyped following the manufacturer's recommendations in three rounds: (i) pilot study including maternal blood and cord blood DNAs, (ii) maternal blood DNAs, (iii) child blood and placenta DNAs. Two HapMap samples were included in each 96-well plate as internal controls of the quality of the whole process. Genotyping clustering was conducted using GenomeStudio GenTrain 3.0. Genetic variants were annotated in b37 + strand using the GSAMD-24v3-0-EA_20034606_A1 manifest.

Quality control of the genome-wide genetic data was conducted with PLINK v1.9 and v2 following the standard recommendations [2]. First, 68,337 genetic variants which were not genotyped in any of the samples were filtered out. Second, we conducted the sample quality control and filtered out samples with a call rate <97% (n=15), discordant for sex (n=12), and with a heterozygosity > +- 5 standard deviations (n=4). Relatedness was estimated using the PI_HAT parameter. All mother-child pairs were identified correctly. Samples with a PI_HAT >0.25, which is indicative of 2nd degree relativeness (n=54) were eliminated, including one of the intended duplicates, mothers participating twice, siblings, other relatives, as well as 16 incorrect samples. Next, we conducted the genetic variant quality control and filtered out SNPs with a call rate <97% (n=14,187), with a minor allele frequency (MAF) <1% (n=151,870), not in Hardy-Weinberg equilibrium (HWE) p-value <1E-06 (n=6,290), or not annotated to any chromosome or with duplicated positions (n=1,277). Ancestry prediction was done using the GRAFpop v2.4 [3]. Predicted ancestries were curated using the questionnaire information on self-reported ethnicity and country of birth of the parents and grandparents, ending up with the following number of participants by main ancestry groups: 1,107 European (76.1%), 327 Latin American (22.5%) and 20 Other (1.4%). We did not filter any sample based on the genetic background. Twenty GWAS principal components (PCs) were estimated for the whole population using a SNP pruning method in the PLINK tool, as well as for the subset of individuals classified as European. The final dataset consisted of 1,454 samples (947 mothers and 507 children, of which 490 were paired).

Up to 40M variants were imputed with the Haplotype Reference Consortium (HRC) version 1.1 panel [4] at the Sanger Institute server (https://www.sanger.ac.uk/tool/sanger-imputation-service), using EAGLE2 for the phasing and PBWT for the imputation. Finally, we removed variants with an imputation quality r2 below 0.9, a MAF lower than 5% and a HWE P-value below 0.05.

**DNA methylation data**

DNAm was assessed in 624 placental DNA samples (including 589 unique individuals and 35 technical duplicates) with the commercial Infinium MethylationEPIC BeadChip from Illumina, following manufacturer's protocol in the Human Genome facility (HUGE193 F) at the Erasmus Medical Centre core facility. The array contains around 850,000 CpG probes, that primarily target gene promoter regions. Additionally, a small number of these probes target FANTOM enhancers and regulatory elements [5]. Briefly, 750 ng of DNA were bisulfite-converted using the EZ 96-DNAm kit following the manufacturer's standard protocol, and DNAm measured using the Infinium protocol.

The methylation data was pre-processed using the PACEAnalysis R package (v.0.1.9), as previously described [6, 7]. The pre-processing pipeline consists of probe quality control, sample quality control, normalization, batch correction, winsorization of outlier values and estimation of cell type proportions. Detection p-values were estimated using out-of-band array hybridization as implemented in the SeSAMe R package [8]. Probe values were masked if the intensity values were zero, estimated based on less than three beads, and/or if they had a detection p-value>0.05. Based on these criteria (low quality (N=5), probe call rate < 95% (N=3), sex inconsistencies (N=9), samples with indication of substantial contamination with maternal DNA or with DNA from another participant in the study (N=11) [9], duplicates identified by clustering samples based on their genetic similarity using the single nucleotide polymorphisms (SNPs) included in the array (N=29) and siblings (N=2). Exclusion of one of the duplicate pairs was done randomly. A total of 565 samples remained after the sample quality control.

After removal of problematic samples, we performed pre-processing of the remaining arrays. Signal intensities were pre-processed by performing linear dye bias correction followed by single-sample background correction based on Normal-exponential convolution using out-of-band Infinium I probes (ssNoob) [10, 11]. Unwanted between-array variation was minimized by applying 220 functional normalization using the control probes [12]. Beta-mixture quantile (BMIQ) normalization was then used to correct for the bias of Type-2 probe values [13]. After that, we explored the clustering of the data through Principal Component Analysis and tested the association of the 12 first principal components (PCs) with technical variables (plate, array, extraction batch, time to placental storage, DNA concentration and 260/280 and 260/230 ratios), design variables (hospital of birth and SARS-CoV-2 confinement) and biological variables (child's sex and ethnic origin). PC1 explained 30.36% of variability, PC2 explained 7.40%, and the rest accounted for less than 3%. The array variable was associated with all the PCs from 1 to 10, and thus, we decided to apply the ComBat method implemented in the sva Bioconductor R package to eliminate its effect [14]. Additionally, sex, ethnicity, gestational age, SARS-CoV-2 confinement and hospital of birth were associated with some of the first PCs.

Following these pre-processing steps, we removed the probes flagged as problematic among the study population. Finally, to correct for the possible outliers, we winsorized the extreme values to the 1% percentile (0.5% in each side), where percentiles were estimated with the empirical beta-distribution. DNAm values are expressed as beta values, where 0 means un-methylation and 1 complete methylation. Final number of probes was 865,859.

Cell type proportions of six main placenta populations (trophoblasts, syncytiotrophoblast, nucleated red blood cell, Hofbauer cells, endothelial cells, and stromal cells) were estimated from DNAm using the reference panel from term placentas implemented in the planet R package [15]. Finally, a DNA contamination score was calculated using SNP probes included in the EPIC array. The score is defined as the average log odds from the SNP posterior probabilities from the outlier component; capturing how irregular the SNP beta-values deviate from the ideal tri-modal distribution [9].

**Placental *cis*-mQTL analysis**

A total of 4,437,007 SNPs and 790,664 CpGs from 408 samples with paired genotype and DNAm data were considered for the *cis*-mQTL analysis in TensorQTL. TensorQTL nominal modality performs linear regressions between the genotype and the normalized DNAm RNT values, as implemented in FastQTL. The *cis*-region was specified as ±0.5Mb from each tested CpG position.

The covariates included in the regression model were the sex of the fetuses, the first five PCs derived from the genotype data (genotype PCs), the first 20 non-genetic DNAm PCs, and the cell type proportions calculated with the Planet methylation panel. Genotype and DNAm PCs were included in the model as covariates to remove hidden and/or technical confounders affecting the DNAm data. To avoid multicollinearity between the non-genetic DNAm PCs and the other covariates, the DNAm PCs were calculated on the residuals from a multiple linear regression adjusting the normalized DNAm RNT-values by the known covariates (sex of the fetuses, the first five genotype PCs, and the six estimated cell types). Following Min et al.[16], we kept all DNAm PCs that cumulatively explained 80% of the variance with a maximum number of 20 PCs for subsequent steps. Then we performed a GWAS on each of the DNAm PCs and retained those PCs that were not associated with the genotype at a suggestive threshold ($P > 1 \times 10^{-7}$). This procedure returned 20 nongenetic DNAm PCs

**Acknowledgement**

We would like to thank all the participants and their families for their generous collaboration. We are also very grateful to all the former and current BiSC team members (https://projectebisc.org/en/team/) for their tremendous contributions to this cohort.

**Funding**

The BiSC cohort has received funding from the European Research Council (ERC) under the European Union's Horizon 2020 research and innovation programme (785994 – AirNB project), from the Health Effects Institute (4959-RFA17-1/18-1 – FRONTIER project), from the European Union's Horizon 2020 research and innovation programme-EU.3.1.2. (874583 - ATHLETE project) and H2020-EU.3.1.1. (GA964827 – AURORA project), from AXA Research Fund (MOOD-COVID project), from Agence nationale de sécurité sanitaire de l'alimentation, de l'environnement et du travail (ANSES) (2019/01/039 - HyPAXE project), from the AGAUR-Agència de Gestió d'Ajuts Universitaris de Recerca (2021 SGR 01570 - Population Neuroscience group), from the Centro de Investigación Biomédica en Red de Epidemiología y Salud Pública (CIBERESP) (CB06/02/0041), from the Instituto de Salud Carlos III (ISCIII) and the European Regional Development Fund (ERDF) - Maternal and Child Health and Development Network (SAMID) (RD16/0022/0014 and RD16/0022/0015), and from the Instituto de Salud Carlos III (ISCIII) and the European Union Next Generation EU - Primary Care Interventions to Prevent Maternal and Child Chronic Diseases of Perinatal and Developmental Origin Network (RICORS-SAMID) (RD21/0012/0001 and RD21/0012/0003). Genome-wide genotyping data was funded by the Instituto de Salud Carlos III (ISCIII) and co-funded by European Union (ERDF) "A way to make Europe" (PI20/01116 – ENTENTE project) and the Centro Nacional de Genotipado-CEGEN (PRB2-ISCIII). Placental DNA methylation was funded by the Instituto de Salud Carlos III (ISCIII) and co-funded by European Union (ERDF) "A way to make Europe" (PI20/00190 – ALMA project), and from the European Joint Programming Initiative "A Healthy Diet for a Healthy Life" (JPI HDHL and Instituto de Salud Carlos III) (AC18/00006 - NutriPROGRAM project). Marta Cosín-Tomàs acknowledges support from the RYC2023-044292-I grant, funded by MICIU/AEI/10.13039/501100011033 and by the FSE+ (this publication is part of the activities supported by this grant). We acknowledge support from the grant CEX2023-0001290-S funded by MCIN/AEI/ 10.13039/501100011033, and support from the Generalitat de Catalunya through the CERCA Program.

#### Early Autism Risk Longitudinal Investigation (EARLI)

**Study Design and Placental Sample**

The Early Autism Risk Longitudinal Investigation (EARLI) is an enriched risk prospective pregnancy cohort to study autism etiology [17]. The EARLI study was reviewed and approved by Human Subjects Institutional Review Boards (IRBs) from each of the four study sites (Johns Hopkins University, Drexel University, University of California Davis, and Kaiser Permanente Northern California). Written informed consent was obtained from all participants. This longitudinal study recruited mothers of confirmed ASD children who were early in a subsequent pregnancy or were trying to become pregnant. There were 232 mothers with a subsequent sibling born through this study. All siblings were born between November 2009 and March 2012.

Placental biopsy samples were collected after delivery at each clinical lab site using Baby Tischler Punch Biopsy Forceps. Paired placental samples in EARLI were taken from the maternal and the child-facing sides of the placenta. Samples were stored at ambient temperature in RNAlater vials (Qiagen) and shipped same-day to the Johns Hopkins Biological Repository (JHBR) in Baltimore, Maryland, for storage at -190°C until DNA processing.

**Genetic Measures**

Genetic data were measured using the Omni5+exome array (Illumina) at the John Hopkins University Center of Inherited Disease Research (CIDR). Data on 4.6 million single nucleotide polymorphisms (SNPs) were generated for 841 EARLI family biosamples (including maternal, paternal, proband, and infant samples) from 254 families and 18 HapMap control samples. Samples were processed together, but only data from infants with placenta methylation data were used. No samples had missing genotypes at >3% of probes, or excess heterozygosity or homozygosity (4 standard deviations). Probes were removed if they had technical problems flagged by CIDR or missing genomic location information. Single nucleotide polymorphisms (SNPs) with minor allele frequencies >5% were removed if they had a missingness rate >5%, and SNPs with minor allele frequency <5% were removed if they had a missingness rate >1%. There were 2.5 million clean SNPs for 827 samples. Imputation was done to the HRC panel on the Michigan Imputation Server.

##### DNA Methylation Measures

Biospecimens including cord blood and placenta were collected and archived at 213 births. Placenta DNA was extracted using the DNA Midi kit (Qiagen, Valencia, CA) and samples were bisulfite treated and cleaned using the EZ DNA methylation gold kit (Zymo Research, Irvine, CA). DNA was plated randomly and assayed on the Infinium HumanMethylation450 BeadChip (Illumina, San Diego, CA) at the Johns Hopkins SNP Center, a shared lab and informatics operation with the Center for Inherited Disease Research (Johns Hopkins University). DNA methylation control gradients and between-plate repeated tissue controls were used.

Quality control of the methylation data was performed using the PACEAnalysis R package (v.0.1.7). Dye-bias and Noob background correction, implemented in the minfi R package, were applied, followed by normalization of the data with the functional normalization method. Then, to correct for the bias of type-2 probe values, the beta-mixture quantile (BMIQ) normalization was applied. To correct for the possible outliers, we Winsorized the extreme values to the 1% percentile (0.5% on each side), where percentiles were estimated with the empirical beta distribution. Finally, the rank-based inverse normal transformation (RNT) was applied to the beta values, and these estimations were the DNAm values considered for mQTL mapping.

Cell type proportions of six populations (STB, TB, nucleated red blood cells, Hofbauer cells, endothelial cells, and stromal cells) were estimated from DNAm using the placenta reference panel from the 3rd trimester implemented in the Planet R package.

##### Placental cis-mQTL analysis

There was n=116 that had both placenta methylation data (431,989 CpGs) and genetic data (4,472,621 SNPs). The cis-mQTL analysis in TensorQTL was done according to consortium specifications. TensorQTL nominal modality performs linear regressions between the genotype and the normalized DNAm RNT values, as implemented in FastQTL. The cis-region was specified as ±0.5Mb from each tested CpG position. Covariates in model included sex, and 5 genetic PCs. In addition, there were 16 methylation PCs after choosing PCs that explained 80% of variance (or maximum of top 20), and excluding PCs strongly associated with genotype.

##### Acknowledgements

We thank the participants of the EARLI study.

##### Funding

Funding for the EARLI study was provided by the National Institutes of Health (R01 ES016443, PI: Newschaffer; R24ES030893, PI: Fallin; R01ES025531, PI: Fallin) and Autism Speaks (003953 PI: Newschaffer). Mr. Dou and Dr. Bakulski were supported by grants from the National Institutes of Health (R01 ES025531, PI: Fallin; R01 MD013299). The content is solely the responsibility of the authors and does not necessarily represent the official views of the National Institutes of Health.

#### Environmental and Childhood Project (Etude des déterminants pré et postnatals du développement de la santé de l'enfant, EDEN)

##### Placental biopsies

EDEN [18] is a population-based, prospective, mother-child cohort study on prenatal and early postnatal nutritional, environmental, and social determinants of the children’s development and health. In total, 2,002 mother-fetus pairs were recruited at the Nancy and Poitiers University hospitals before their 24th week of gestation.

##### Genotype data

DNA was extracted from cord blood samples. Genome-wide genotyping was performed using the Illumina Global Screening Array chip (650k). A quality control [19] process was applied on variants and individuals using PLINK (version 2.00a6LM of the 14 April 2024). After this filtering step, 1,363 women; and approximately 600,000 SNPs remained.

The quality control of the genotype data from 1,004 EDEN samples and 500,689 genetic variants was performed using the PLINK 1.9 software following the standard recommendations. All plink files were initially processed with Will Rayner's preparation Perl script available from Mark McCarthy's Group as recommended in the documentation from the Michigan Imputation Server, using the HRC r1.1 2016 reference panel. Variants with a call rate below 95%, minor allele frequency (MAF) below 1%, or a P-value from the Hardy-Weinberg Equilibrium (HWE) exact test below $1 \times 10^{-6}$ were removed. Samples with discordant sex, those with average heterozygosity values above or below 4 standard deviations, or with more than 3% missing genotype were filtered out. Identity-by-descent values were calculated with PLINK, and from those sample pairs that showed PI-HAT estimates above 0.18, the sample with a higher proportion of missing genotypes was removed.

The final dataset was imputed with the Michigan Imputation Server using the 1000 Genomes phase 3, Version 2.0.0. Before imputation, data was converted into VCF format. Phasing of haplotypes was done with Eagle v2.4 and genotype imputation with Minimac4, both implemented in the code by the Michigan Imputation Server. Finally, we removed variants with an imputation quality $r^2$ below 0.9, a MAF lower than 5% and a HWE P-value below 0.05. Only those samples with paired DNAm data were considered in this analysis. The final dataset consisted of 976 samples and 4,125,333 SNPs.

**DNA extraction and DNA methylation data**

At delivery, placental tissue from the fetal side was sampled at one site by the specifically-trained midwives of the study using the following standardized procedure. Samples of around $5mm^3$ were collected a few centimeters from the insertion of the cord under the chorio-amniotic membrane, washed in a saline solution and immediately frozen at -80°C. The protocol was similar for both centers and all modes of delivery.

For 668 EDEN samples, DNA was extracted using the QIAsymphony instrument (Qiagen, Germany) and whole-genome DNAm was measured using the Infinium HumanMethylation450 BeadChip. An additional set of 382 EDEN (EPIMEX) samples was processed with the DNeasy Blood & Tissue Kit (Qiagen) using the DNeasy® Blood & Tissue Handbook according to the manufacturer's instructions and whole-genome DNAm was measured using the Infinium HumanMethylationEPIC *BeadChip*. Raw intensities of fluorescent signals were extracted using the GenomeStudio→ software. DNA samples were allocated so that the ratios for sex (boy/girl) and recruitment center (Poitiers/Nancy) were balanced for each chip.

The quality control of the DNAm data, including 866,553 and 486,427 DNAm probes from EPIC and 450k, respectively, was performed using the PACEAnalysis R package (v.0.1.7). Before the quality control with PACEAnalysis, one sample was discarded because of too many missing values in relevant variables. With the R package, we discarded those samples with a call rate below 95%, sex inconsistencies, intentioned or non-intentioned duplicates, and those contaminated with DNA from another subject or the mother.

Only those samples with paired genotype data were considered in this study. Probes with a call rate lower than 95% and in the sex chromosomes were excluded from the analysis.

The methylation beta values were normalized in different steps. Dye-bias and Noob background correction, implemented in the minfi R package, were applied, followed by normalization of the data with the functional normalization method. Then, to correct for the bias of type-2 probe values, the beta-mixture quantile (BMIQ) normalization was applied. After that, we explored the clustering of the data through Principal Component Analysis (PCA) and tested the association of the 12 first PCs with the main and the technical variables. Array batch effect was controlled with the ComBat method. To correct for the possible outliers, we Winsorized the extreme values to the 1% percentile (0.5% on each side), where percentiles were estimated with the empirical beta distribution. Finally, the rank-based inverse normal transformation (RNT) was applied to the beta values, and these estimations were the DNAm values considered for mQTL mapping. The final dataset consisted of 375 and 659 samples and 708,802 and 441,073 DNAm probes (CpGs) from EPIC and 450k, respectively.

Cell type proportions of six populations (STB, TB, nucleated red blood cells, Hofbauer cells, endothelial cells, and stromal cells) were estimated from DNAm using the placenta reference panel from the 3$^{rd}$ trimester implemented in the Planet R package.

**Placental cis-mQTL analysis in EPIC**

A total of 4,125,333 SNPs and 708,802 CpGs from 203 samples with paired genotype and DNAm data were considered for the *cis*-mQTL analysis in TensorQTL. TensorQTL nominal modality performs linear regressions between the genotype and the normalized DNAm RNT values, as implemented in FastQTL. The *cis*-region was specified as ±0.5Mb from each tested CpG position.

The covariates included in the regression model were the sex of the fetuses, the first five PCs derived from the genotype data (genotype PCs), the first 19 non-genetic DNAm PCs, and the cell type proportions calculated with the Planet methylation panel. Genotype and DNAm PCs were included in the model as covariates to remove hidden and/or technical confounders affecting the DNAm data. To avoid multicollinearity between the non-genetic DNAm PCs and the other covariates, the DNAm PCs were calculated on the residuals from a multiple linear regression adjusting the normalized DNAm RNT-values by the known covariates (sex of the fetuses, the first five genotype PCs, and the five estimated cell types). Following Min et al., we kept all DNAm PCs that cumulatively explained 80% of the variance with a maximum number of 20 PCs for subsequent steps. Then we performed a GWAS on each of the DNAm PCs and retained those PCs that were not associated with the genotype at a suggestive threshold ($P > 1 \times 10^{-7}$). This procedure returned 19 non-genetic DNAm PCs.

**Placental cis-mQTL analysis in 450k**

A total of 4,125,333 SNPs and 441,073 CpGs from 329 samples with paired genotype and DNAm data were considered for the *cis*-mQTL analysis in TensorQTL. TensorQTL nominal modality performs linear regressions between the genotype and the normalized DNAm RNT values, as implemented in FastQTL. The *cis*-region was specified as ±0.5Mb from each tested CpG position.

The covariates included in the regression model were the sex of the fetuses, the first five PCs derived from the genotype data (genotype PCs), the first 20 non-genetic DNAm PCs, and the cell type proportions calculated with the Planet methylation panel. Genotype and DNAm PCs were included in the model as covariates to remove hidden and/or technical confounders affecting the DNAm data. To avoid multicollinearity between the non-genetic DNAm PCs and the other covariates, the DNAm PCs were calculated on the residuals from a multiple linear regression adjusting the normalized DNAm RNT-values by the known covariates (sex of the fetuses, the first five genotype PCs, and the five estimated cell types). Following Min et al., we kept all DNAm PCs that cumulatively explained 80% of the variance with a maximum number of 20 PCs for subsequent steps. Then we performed a GWAS on each of the DNAm PCs and retained those PCs that were not associated with the genotype at a suggestive threshold ($P > 1 \times 10^{-7}$). This procedure returned 20 non-genetic DNAm PCs.

**Acknowledgement**

We are extremely grateful to all the families who took part in this study, the midwives and psychologists for recruiting and following them, and the whole EDEN team, including research scientists, engineers, technicians and managers for their commitment and their role in the success of the study.

**Funding**

The EDEN cohort has been funded by the Foundation for Medical Research (FRM), National Agency for Research (ANR), National Institute for Research in Public Health (IRESP: TGIR cohorte santé 2008 program), French Ministry of Health (DGS), French Ministry of Research, Paris–Sud University, French National Institute for Population Health Surveillance (Santé Publique France, ex InVS), the European Union FP7 programs (FP7/2007-2013), French Agencies for Environmental Health and Food Safety (now ANSES), Mutuelle Générale de l'Education Nationale (MGEN), and the French Society for Diabetes. L.B. was supported by a grant from the French Agency for National Research (grant number ETAPE, ANR-18-CE36–0005) and the Fondation pour la Recherche Médicale (EPImEx project). EDEN DNA methylation measurements were obtained thanks to grants from the Fondation de France (grant numbers 2012–00031593, 2012–00031617) and the Fondation pour la Recherche Médicale (EPImEx project).

#### Genetics of Glucose regulation in Gestation and Growth (Gen3G) cohort

##### Placental biopsies and DNA extraction

The Gen3G prospective cohort recruited 1,024 pregnant women during their first trimester of pregnancy in Sherbrooke, Canada. Immediately after delivery, research staff collected umbilical cord blood and placenta samples. Placental samples (1 cm3) were obtained approximately 5 cm from the umbilical cord insertion from the fetal side and stored in RNAlater (Qiagen) at -80°C until DNA extraction. DNA was purified using the AllPrep DNA Mini Kit (Qiagen), and purity was assessed by spectrophotometry (Ultrospec 2000 UV/Visible; Pharmacia Biotech) using the 260/280 nm absorbance ratio, as recommended [20]. DNA from the cord blood samples was isolated using the Gentra Puregene Blood Kit (Qiagen, Mississauga, ON, Canada).

##### Genotype data

Illumina short-read whole genome sequencing (WGS) was performed on 471 samples using 400 ng of umbilical cord blood DNA and the NxSeq AmpFREE low DNA Library Kit (Lucigen, LGC, UK) according to manufacturer's recommendations. Libraries were sequenced on an Illumina HiSeqX instrument targeting 16x coverage at two libraries per flowcell lane. Resulting data were processed with the DNA-Seq v3.1.4 pipeline from GenPipes [21] based on BWA_mem [22] and GATK 3.8 [23] best practices (omitting base requalibration) using GRCh37 for alignment. Identification of high-quality SNPs was done through joint genotyping over all samples [24]. Samples in the resulting VCF with a high fraction of heterozygous sites (heterozygocity estimated with Plink 1.9) were considered contaminated and removed (N = 3).

The VCFs were lifted over to GRCh38 using the tool LiftoverVcf version 2.27.4 from Picard Tools [25]. Imputation of missing data in our WGS dataset was conducted using GLIMPSE2 [26]. The reference panel for imputation was prepared using GLIMPSE2_chunk and GLIMPSE2_split_reference on the high-coverage expanded 1000 Genome Project dataset [27]. Imputation was then performed using GLIMPSE2_phase. Imputed and sequenced data were merged using a custom Python script that only added data from the imputed VCF to the lifted VCF if it was missing in the latter (eg. missing genotypes and variants). Variants were annotated using dbSNP v156 [28]. Only SNPs with a minor allele frequency above 5% and a Hardy-Weinberg Equilibrium exact test p-value above 0.05 were kept. After filtering samples to keep only individuals with placental DNAm data, the final dataset contained 408 individuals and 6,040,431 SNPs.

##### DNA methylation data

DNAm was assessed with the Infinium Methylation EPIC BeadChip from Illumina, following the manufacturer's protocol. Briefly, 750 ng of DNA from 524 placental samples were bisulfite-converted using the EZ 96-DNA methylation kit from Zymo Research, following the manufacturer's standard protocol, and DNAm was measured using the Infinium protocol. Fifteen technical duplicates were included initially for quality control procedures.

The quality control of the DNAm data, initially from 865,859 DNAm probes, was performed using the PACEAnalysis R package (v.0.1.7). With the R package, we excluded samples with a call rate below 95%, sex inconsistencies, intentioned or non-intentioned duplicates, and those contaminated with DNA from another subject or the mother. Probes with a call rate lower than 95% and in the sex chromosomes were excluded from the analysis.

The methylation beta values were normalized in different steps. Dye-bias and Noob background correction, implemented in the minfi R package, were applied, followed by normalization of the data with the functional normalization method. Then, to correct for the bias of type-2 probe values, the beta-mixture quantile (BMIQ) normalization was applied. After that, we explored the clustering of the data through Principal Component Analysis (PCA) and tested the association of the 12 first PCs with the main and the technical variables. Array batch effect was controlled with the ComBat method. To correct for the possible outliers, we Winsorized the extreme values to the 1% percentile (0.5% on each side), where percentiles were estimated with the empirical beta distribution. Cell type proportions of six populations (syncytiotrophoblast, trophoblast, nucleated red blood cells, Hofbauer cells, endothelial cells, and stromal cells) were estimated from DNAm using the placenta reference panel from the 3rd trimester implemented in the Planet R package. Four samples with low syncytiotrophoblast proportion (< 25%) were removed. Finally, the rank-based inverse normal transformation (RNT) was applied to the beta values, and these estimations were the DNAm values considered for mQTL mapping. The final dataset consisted of 408 placenta samples and 765,790 DNAm probes (CpGs).

**Placental *cis*-mQTL analysis**

A total of 6,040,431 SNPs and 765,790 CpGs from 408 samples with paired genotype and placental DNA methylation data were included in the cis-mQTL analysis using TensorQTL. The nominal mode in TensorQTL, as implemented in FastQTL, performs linear regressions between genotypes and normalized DNA methylation (RNT) values. *Cis*-regions were defined as ±0.5 Mb from each CpG site.

Covariates included in the regression model were fetal sex, the first five genotype principal components (PCs), 20 non-genetic DNA methylation (DNAm) PCs, and cell type proportions estimated using the Planet methylation panel. Genotype and DNAm PCs were included to account for hidden and technical confounders affecting DNAm data. To avoid multicollinearity, DNAm PCs were computed on residuals obtained from a multiple linear regression of normalized DNAm RNT values adjusted for known covariates (fetal sex, the first five genotype PCs, and five estimated cell types). Following the approach of Min et al., DNAm PCs cumulatively explaining up to 80% of the variance (maximum of 20 PCs) were initially selected. Subsequently, genome-wide association studies (GWAS) were performed on each DNAm PC, and PCs showing no association with genotype at a suggestive threshold ($P > 1 \times 10^{-7}$) were retained. This procedure resulted in 20 non-genetic DNAm PCs used in the final analysis.

**Acknowledgement**

We thank all Gen3G participants for their time and involvement in the Gen3G cohort.

**Funding**

Gen3G was supported over the years by the Fonds de Recherche du Québec en Santé (FRQ-S Grant #20697), Canadian Institute of Health Research (CIHR) (Grants #MOP115071, #PJT152989 and #PJT190076) operating grants, Diabetes Quebec, and an American Diabetes Association (ADA) Pathways To Stop Diabetes Accelerator Award (#1-15-ACE-26). LB and PEJ are senior research scholars from the FRQS. The DNA sequencing of the Gen3G offspring has been supported by Fonds de recherche du Québec, McGill University and Université de Sherbrooke.

### Environmental and Childhood Project (INfancia y Medio Ambiente, INMA)

**Placental biopsies and DNA extraction**

In the INMA project [29], 2,506 mother-fetus pairs were followed until birth, and a selection of 397 placentas were collected, representing the three geographical areas involved in the study. Collected placentas were stored at −80 °C in a central biobank until processing. Biopsies of approximately 5 $cm^3$ were obtained from the inner region of the placenta, approximately 1.0–1.5 cm below the fetal membranes, corresponding to the villous parenchyma, and at a distance of ~5 cm from the site of cord insertion. 25mg of placental tissue was used for DNA extraction, previously rinsed twice for 5min in 0.8mL of 0.5X PBS in order to remove traces of maternal blood. Genomic DNA from the placenta was isolated using the DNAeasy® Blood and Tissue Kit (Qiagen, CA, USA). DNA quality was evaluated on a NanoDrop spectrophotomer (Thermo Scientific, Waltham, MA, USA) and additionally, 100 ng of DNA was run on 1.3% agarose gels to confirm that samples did not present visual signs of degradation. Isolated genomic DNA was stored at −20 °C until further processing.

**Genotype data**

Genome-wide genotyping was performed using the Illumina GSA Beadchip at the Human Genotyping Facility (HuGeF), Dept Internal Medicine, Erasmus MC, Rotterdam, the Netherlands, and the Spanish National Genotyping Center, CEGEN, Madrid, Spain. Genotype calling was done using the GeneTrain2.0 algorithm based on HapMap clusters implemented in the Genome Studio software. Samples were genotyped in four batches.

The quality control of the genotype data from 397 INMA samples and 509,450 genetic variants was performed using the PLINK 1.9 software following the standard recommendations. All plink files were initially processed with Will Rayner's preparation Perl script available from Mark McCarthy's Group as recommended in the documentation from the Michigan Imputation Server, using the HRC r1.1 2016 reference panel. Variants with a call rate below 95%, minor allele frequency (MAF) below 1%, or a P-value from the Hardy-Weinberg Equilibrium (HWE) exact test below $1 \times 10^{-6}$ were removed. Samples with discordant sex, those with average heterozygosity values above or below 4 standard deviations, or with more than 3% missing genotype were filtered out. Identity-by-descent values were calculated with PLINK, and from those sample pairs that showed PI-HAT estimates above 0.18, the sample with a higher proportion of missing genotypes was removed.

The final dataset was imputed with the Michigan Imputation Server using the HRC reference panel, Version r.1.1 2016. Before imputation, data was converted into VCF format. Phasing of haplotypes was done with Eagle v2.4 and genotype imputation with Minimac4, both implemented in the code by the Michigan Imputation Server. Finally, we removed variants with an imputation quality $r^2$ below 0.9, a MAF lower than 5% and a HWE P-value below 0.05. Only those samples with paired DNAm data were considered in this analysis. The final dataset consisted of 368 samples and 4,171,035 SNPs.

**DNA methylation data**

DNAm was assessed with the Infinium Methylation EPIC BeadChip from Illumina, following the manufacturer's protocol in the Erasmus Medical Center core facility. Briefly, 750 ng of DNA from 397 placental samples were bisulfite-converted using the EZ 96-DNA methylation kit from Zymo Research, following the manufacturer's standard protocol, and DNAm was measured using the Infinium protocol. Three technical duplicates were included. Samples were randomized taking into account region-of-origin and sex. As the number of samples in each condition was different, a perfect randomization was not possible.

However, all the plates had samples from all three geographical areas involved, and an equilibrated number of male and female samples. The quality control of the DNAm data, including 865,859 DNAm probes, was performed using the PACEAnalysis R package (v.0.1.7). Before the quality control with PACEAnalysis, one sample was discarded because of too many missing values in relevant variables. With the R package, we discarded those samples with a call rate below 95%, sex inconsistencies, intentioned or non-intentioned duplicates, and those contaminated with DNA from another subject or the mother.

Only those samples with paired genotype data were considered in this study. Probes with a call rate lower than 95% and in the sex chromosomes were excluded from the analysis.

The methylation beta values were normalized in different steps. Dye-bias and Noob background correction, implemented in the minfi R package, were applied, followed by normalization of the data with the functional normalization method. Then, to correct for the bias of type-2 probe values, the beta-mixture quantile (BMIQ) normalization was applied. After that, we explored the clustering of the data through Principal Component Analysis (PCA) and tested the association of the 12 first PCs with the main and the technical variables. Array batch effect was controlled with the ComBat method. To correct for the possible outliers, we Winsorized the extreme values to the 1% percentile (0.5% on each side), where percentiles were estimated with the empirical beta distribution. Finally, the rank-based inverse normal transformation (RNT) was applied to the beta values, and these estimations were the DNAm values considered for mQTL mapping. The final dataset consisted of 368 samples and 795,055 DNAm probes (CpGs).

Cell type proportions of six populations (STB, TB, nucleated red blood cells, Hofbauer cells, endothelial cells, and stromal cells) were estimated from DNAm using the placenta reference panel from the 3$^{rd}$ trimester implemented in the Planet R package.

**Placental *cis*-mQTL analysis**

A total of 4,171,035 SNPs and 795,055 CpGs from 368 samples with paired genotype and DNAm data were considered for the *cis*-mQTL analysis in TensorQTL. TensorQTL nominal modality performs linear regressions between the genotype and the normalized DNAm RNT values, as implemented in FastQTL. The *cis*-region was specified as ±0.5Mb from each tested CpG position.

The covariates included in the regression model were the sex of the fetuses, the first five PCs derived from the genotype data (genotype PCs), the first 18 non-genetic DNAm PCs, and the cell type proportions calculated with the Planet methylation panel. Genotype and DNAm PCs were included in the model as covariates to remove hidden and/or technical confounders affecting the DNAm data. To avoid multicollinearity between the non-genetic DNAm PCs and the other covariates, the DNAm PCs were calculated on the residuals from a multiple linear regression adjusting the normalized DNAm RNT-values by the known covariates (sex of the fetuses, the first five genotype PCs, and the five estimated cell types). Following Min et al., we kept all DNAm PCs that cumulatively explained 80% of the variance with a maximum number of 20 PCs for subsequent steps. Then we performed a GWAS on each of the DNAm PCs and retained those PCs that were not associated with the genotype at a suggestive threshold ($P > 1 \times 10^{-7}$). This procedure returned 18 nongenetic DNAm PCs.

**Acknowledgement**

We thank all the participants and their families.

**Funding**

INMA-Gipuzkoa is funded by grants from Instituto de Salud Carlos III (PI06/0867 and PI09/00090, incl. FEDER funds), Department of Health of the Basque Government (2005111093), Provincial Government of Gipuzkoa (DFG06/002), and annual agreements with the municipalities of the study area (Zumarraga, Urretxu, Legazpi, Azkoitia, Azpeitia and Beasain). INMA-Sabadell was funded by grants from Instituto de Salud Carlos III (Red INMA G03/176, PS09/00432, PI17/01225, PI17/01935, and CP18/00018), Fundació La Marató de TV3 (090430), and Generalitat de Catalunya-CIRIT (1999SGR 00241), the European Community's Seventh Framework Program (FP7/2007-206) under grant agreement no 308333 (HELIX project), and from the European Joint Programming

Initiative “A Healthy Diet for a Healthy Life” (JPI HDHL and Instituto de Salud Carlos III) under the grant agreement no AC18/00006 (NutriPROGRAM project). ISGlobal acknowledges support from the Spanish Ministry of Science and Innovation and the State Research Agency through the “Centro de Excelencia Severo Ochoa 2019-2023” Program (CEX2018-000806-S), and support from the Generalitat de Catalunya through the CERCA Program. INMA-Valencia is funded by Grants from UE (FP7-ENV-2011 cod 282957, HEALTH.2010.2.4.5-1, and H2020 No 874583, the ATHLETE project), the Ministry of Universities (CAS21/00008, Margarita Salas Grant MS21-133 and NextGeneration EU), Instituto de Salud Carlos III (FIS-FEDER: 13/1944, 16/1288, 17/00663, and 19/1338; FIS-FSE: 17/00260; Miguel Servet-FSE: MSII20/0006; PI23/01578; PFIS: FI24/00055), CIBERESP, Generalitat Valenciana (CIGE23/142, CIAICO/2021/132, BEST/2020/059, and AICO 2020/285). M.C.-T. is funded by a Beatriu de Pinós Postdoctoral Contract awarded by Generalitat de Catalunya-AGAUR and European Commission- Horizon 2020 (2019 BP 00107). M.F.F. is funded by the EU Commission (QLK4-1999-01422, QLK4-2002-00603, and CONTAMED FP7-ENV-212502) and the Consejería de Salud de la Junta de Andalucía (Grant number 0675/10). J.R.B. is funded by Research Grant PID2019-106382RB-I00 funded by MCIN/AEI/10.13039/501100011033. I.G.M. is funded by the JDC2023-051497-I research grant from the MCIU/AEI/10.13039/501100011033 and the FSE+. N.F.J. is funded by research grants 2024111079 from the Basque Department of Health, EHU-G24/06 from the University of the Basque Country (EHU), and PI21/01491 from ISCIII, co-funded by the European Union.

### InTraUterine sample in early pregnancy (ITU)

##### Study design

The Intrauterine Sampling in Early Pregnancy (ITU) is a cohort comprising 943 pregnant Finnish women and their children born between 2012 and 2017. Recruitment took place through the national voluntary prenatal screening program for trisomy 21 at maternity clinics in Helsinki and Uusimaa Hospital District in Finland. The ITU study comprises two study arms: women in the chromosomal testing arm (N=544) had an increased risk of fetal chromosomal abnormalities based on the screening program (routine serum, ultrasound, age, maternal height and weight, patient history) and were offered fetal chromosomal testing including CVS at the Helsinki and Uusimaa Hospital District Fetomaternal Medical Center. However, only women with a negative testing result (i.e., no fetal chromosomal abnormalities) were contacted for final recruitment. Women in the no-chromosomal-testing arm (N=399) were informed about the study when attending the same routine serum and ultrasound screening but had no increased risk for fetal chromosomal abnormalities and thus no chromosomal testing. For all women, eligibility criteria included singleton pregnancy, no prenatal diagnosis of chromosomal abnormality, maternal age ≥18 years, and sufficient Finnish language ability to ensure informed consent.

The ITU research protocol has been approved by the Coordinating Ethics Committee of the Helsinki and Uusimaa Hospital District (approval date, 18 May 2010; reference number, 269/13/03/00/09). The study protocol follows the Helsinki Declaration. Participants provided written informed consent to participate in the study before taking part.

##### Placenta tissue samples

First-trimester placenta tissue samples were available from leftover CVS, taking place between 10 and 15 gestational weeks. Birth placenta samples were collected with nine-site biopsies from the fetal side of the placenta (at 2–3 cm from umbilical cord insertion) within maximum 120 min after delivery and stored in RNA storage solution (RNAprotect, Qiagen). Tissue biopsies were stored at −80°C. Quantification and quality assessments were performed using a TapeStation Automated Electrophoresis system (Agilent) and an Epoch Microplate Spectrophotometer (BioTek, Agilent). All extractions were performed at the BioPrep core unit, Max Planck Institute of Psychiatry.

##### Genotyping

Genotyping was performed on Illumina GSA-24v2-0_A1 arrays according to the manufacturer's guidelines. Quality control (QC) was performed in plink v1.9 and R using a standard QC pipeline. SNPs with a minor allele frequency (MAF) below 1%, a call rate below 98%, or with deviation from Hardy-Weinberg equilibrium (HWE) with a p value $< 1 \times 10^{-5}$ were removed from the analysis. Furthermore, we removed SNPs mapping to multiple locations as well as duplicated variants. Individuals with a genotype call rate below 98% were excluded. We identified seven sample pairs with identity-by-descent estimates >0.125, one sample was removed from each pair. No individuals showed discrepancies between phenotypic and genotypic sex. After the first QC, the dataset comprised 338,132 SNPs and 592 individuals. Genotypes were imputed using shapeit2 impute2 plink v1.9, and plink2. Chromosomal and base pair positions were updated to the 1000 Genomes Phase 3 reference set, and allele strands were flipped where necessary. Then haplotype phasing, followed by imputation, was performed. Afterwards, SNPs with an info score <0.6, MAF below 1%, and a deviation from HWE with a p value $<1 \times 10^{-5}$ were filtered out. Finally, genotypes were converted into best-guessed genotypes with plink and a hard-call threshold of 0.1. This resulted in a dataset of 9,774,761 SNPs for 592 individuals. Afterwards, an additional QC was performed on the best-guessed genotypes filtering for hard-called SNP genotypes with MAF < 0.05, call rate <0.98, and HWE $p <1 \times 10^{-5}$. This led to a final 3,513,721 SNPs among 592 individuals.

To retrieve ancestry-related information, we performed multi-dimensional scaling (MDS) analysis on the identity-by-state matrix of quality-controlled genotypes after linkage disequilibrium pruning with a step size of 50, a 250-kb window, and $r^2 = 0.2$. Outliers, defined as samples presenting with a position on any of the first 10 axes of variation deviating more than four standard deviations from the respective axis' mean, were iteratively removed until no more outliers were detected. Afterwards, individuals presenting with heterozygosity values more than four standard deviations away from the mean heterozygosity were also iteratively removed (n = 2). The first two MDS components were extracted.

**Placental methylation**

The quality control of the DNAm data was performed using the PACEAnalysis R package (v.0.1.7 ; https://www.epicenteredresearch.com/ ). With the R package, we discarded those samples with a call rate below 95%, sex inconsistencies, intentioned or non-intentioned duplicates, and those contaminated with DNA from another subject or the mother.

Only those samples with paired genotype data were considered in this study. Probes with a call rate lower than 95% and in the sex chromosomes were excluded from the analysis.

The methylation beta values were normalized in different steps. Dye-bias and Noob background correction, implemented in the minfi R package, were applied, followed by normalization of the data with the functional normalization method. Then, to correct for the bias of type-2 probe values, the beta-mixture quantile (BMIQ) normalization was applied. After that, we explored the clustering of the data through Principal Component Analysis (PCA) and tested the association of the 12 first PCs with the main and the technical variables. Array batch effect was controlled with the ComBat method. To correct for the possible outliers, we Winsorized the extreme values to the 1% percentile (0.5% on each side), where percentiles were estimated with the empirical beta distribution. Finally, the rank-based inverse normal transformation (RNT) was applied to the beta values, and these estimations were the DNAm values considered for mQTL mapping. The final dataset consisted of 392 samples and 669,776 DNAm probes (CpGs).

Cell type proportions of six populations (STB, TB, nucleated red blood cells, Hofbauer cells, endothelial cells, and stromal cells) were estimated from DNAm using the placenta reference panel from the 3rd trimester implemented in the Planet R package.

**Placental *cis*-mQTL analysis**

A total of 6,474,180 SNPs and 669,776 CpGs from 392 samples with paired genotype and DNAm data were considered for the *cis*-mQTL analysis in TensorQTL. TensorQTL nominal modality performs linear regressions between the genotype and the normalized DNAm RNT values, as implemented in FastQTL. The *cis*-region was specified as ±0.5Mb from each tested CpG position.

The covariates included in the regression model were the sex of the fetuses, the first five PCs derived from the genotype data (genotype PCs), the first 18 non-genetic DNAm PCs, and the cell type proportions calculated with the Planet methylation panel. Genotype and DNAm PCs were included in the model as covariates to remove hidden and/or technical confounders affecting the DNAm data. To avoid multicollinearity between the non-genetic DNAm PCs and the other covariates, the DNAm PCs were calculated on the residuals from a multiple linear regression adjusting the normalized DNAm RNT-values by the known covariates (sex of the fetuses, the first five genotype PCs, and the five estimated cell types). Following Min et al., we kept all DNAm PCs that cumulatively explained 80% of the variance with a maximum number of 20 PCs for subsequent steps. Then we performed a GWAS on each of the DNAm PCs and retained those PCs that were not associated with the genotype at a suggestive threshold ($P > 1 \times 10^{-7}$). This procedure returned 18 nongenetic DNAm PCs.

**Funding**

The ITU study is funded by the Research Council of Finland (award numbers: 1284859, 12848591, 312670, 1324596) and the Diabetes Research Foundation.

**Acknowledgements**

We thank all the participants of the ITU study and all the study nurses, especially Eija Lahdensuo for the sample and data collection.

#### Markers of Autism Risk Learning Early Signs (MARBLES)

**Study Design and Placental Sample**

Markers of Autism Risk Learning Early Signs (MARBLES) is an enriched risk prospective pregnancy cohort to study autism etiology [30]. The MARBLES protocol was reviewed and approved by the Human Subjects Institutional Review Board (IRB) from University of California Davis. Written informed consent was obtained from all participants. This ongoing longitudinal study recruited mothers of confirmed ASD children who were in a subsequent pregnancy or were trying to become pregnant. At the time of these analyses, there were 389 enrolled mothers that gave birth to 425 subsequent siblings between December 1, 2006 and July 1, 2016.

At the delivery hospital, placental tissues are collected and immediately processed and frozen. Because human term placenta is large and heterogeneous, the MARBLES study used orientation to the umbilical cord to ensure that all samples are isolated from a similar region, the chorionic villus from the fetal side of the placenta. Placental samples were taken from the child-facing sides of the placenta and stored at -80 degrees Celsius in the UC Davis repository.

**Genetic Measures**

In MARBLES, SNPs on 643 infant and mother samples from 234 families were genotyped using the Illumina Mega array at the John Hopkins University Center of Inherited Disease Research (CIDR). Maternal and infant samples were processed together, but only data from infants with placenta methylation measures were used. We applied stringent quality control criteria to the raw 1.75 million genotypes to remove low quality SNPs and samples. Our criteria include removal of samples with call rate <98%, sex discrepancy, and relatedness (pi-hat < 0.18) to non-familial samples. We also filtered SNPs with call rates < 95%, excess hetero- or homozygosity, and minor allele frequency (MAF) < 5%. After quality control, 620 samples and 758 thousand SNPs remained. Imputation was done to the HRC panel on the Michigan Imputation Server.

**DNA Methylation Measures**

Placenta biospecimens were collected and archived. Biopsy punch (<5 mg) of frozen placenta tissue were transferred to the LaSalle laboratory where placenta DNA was extracted from a subsample of 92 participants with 36-month diagnostic assessment data available using the DNA Midi kit (Qiagen, Valencia, CA). Samples were bisulfite treated and cleaned using the EZ DNA methylation gold kit (Zymo Research, Irvine, CA). DNA was plated randomly and assayed on the Infinium HumanMethylationEPIC BeadChip (Illumina, San Diego, CA) at the Johns Hopkins SNP Center, a shared lab and informatics operation with the Center for Inherited Disease Research (Johns Hopkins University).

Quality control of the methylation data was performed using the PACEAnalysis R package (v.0.1.7). Dye-bias and Noob background correction, implemented in the minfi R package, were applied, followed by normalization of the data with the functional normalization method. Then, to correct for the bias of type-2 probe values, the beta-mixture quantile (BMIQ) normalization was applied. To correct for the possible outliers, we Winsorized the extreme values to the 1% percentile (0.5% on each side), where percentiles were estimated with the empirical beta distribution. Finally, the rank-based inverse normal transformation (RNT) was applied to the beta values, and these estimations were the DNAm values considered for mQTL mapping.

Cell type proportions of six populations (STB, TB, nucleated red blood cells, Hofbauer cells, endothelial cells, and stromal cells) were estimated from DNAm using the placenta reference panel from the 3rd trimester implemented in the Planet R package.

**Placental cis-mQTL analysis**

There was n=75 that had both placenta methylation data (814,590 CpGs) and genetic data (4,608,539 SNPs). The cis-mQTL analysis in TensorQTL was done according to consortium specifications. TensorQTL nominal modality performs linear regressions between the genotype and the normalized DNAm RNT values, as implemented in FastQTL. The *cis*-region was specified as ±0.5Mb from each tested CpG position. Covariates in model included sex, and 5 genetic PCs. In addition, there were 4 methylation PCs after choosing PCs that explained 80% of variance (or maximum of top 20), and excluding PCs strongly associated with genotype.

**Acknowledgements**

We are grateful to the participants of the MARBLES Study, without whom this work would not have been possible.

**Funding Information**

The MARBLES study and this work has been supported by a grant from the Allen Foundation, pilot funding from the MIND Institute, EPA STAR grant #RD-83329201, and NIH grants: R24ES028533, R01ES028089, R01ES020392, R01ES025574, P01ES011269, and K12HD051958. These supporting organizations had no role in the design and conduct of the work; collection, management, analysis, and interpretation of the data; preparation, review, or approval of the manuscript; and decision to submit the manuscript for publication. The findings and conclusions in this report are those of the authors and do not necessarily represent the official position of the National Institutes of Health or EPA.

### The Prediction and Prevention of Preeclampsia and Intrauterine Growth Restriction (PREDO)

#### Study design

The Prediction and Prevention of Preeclampsia and Intrauterine Growth Restriction (PREDO) is a prospective birth cohort study of Finnish women who were pregnant between 2005 and 2010 and their children. The PREDO study cohort was set up to identify novel risk factors and biomarkers in pregnant women associated with the development of preeclampsia and intrauterine growth restriction (IUGR), to (a) identify effective methods for prediction and prevention of preeclampsia in at-risk women, and (b) determine the association between exposure to preeclampsia, IUGR, or their risk factors and child developmental/health outcomes. Women with a singleton, intrauterine pregnancy who visited antenatal clinics at ten study hospitals in Finland for their first ultrasound screening at 12+0-13+6 weeks+days of gestation were recruited in the PREDO study. Two groups of pregnant women were enrolled: first, pregnant women with a known clinical risk factor status for preeclampsia and IUGR, and second, pregnant women who volunteered to participate regardless of their risk factor status for preeclampsia and IUGR. The sample with a known risk factor status comprises 1,079 pregnant women who gave live birth (969 of these women had at least one and 110 had none of the known risk factors for preeclampsia and IUGR). Of the sample, N=140 placenta nine-site biopsies were taken from the decidual side of the placenta maximum of 90 mins after delivery.

The study protocol was approved by the Ethics Committee of Obstetrics and Gynaecology, and Women, Children and Psychiatry of the Helsinki and Uusimaa Hospital District and by the participating hospitals (Dnro 5/E8/05, Dnro 3/E8/05, 216/13/03/03/2012). All participants provided written informed consent. Consent of participating children were provided by parent(s)/guardian(s). Details of the study design, inclusion criteria, enrollment and data collection are described elsewhere (1). The study has been registered as ClinicalTrials.gov identifier ISRCTN14030412.

In the high-risk sample, blood samples in the mothers and fathers and cord blood samples were collected according to standard procedures. DNA was extracted at the National Institute for Health and Welfare, Helsinki, Finland and the Finnish Institute of Molecular Medicine, University of Helsinki, Finland and methylation analyses were performed at the Max Planck Institute in Munich, Germany.

#### Genotyping

Genotyping was performed on the IlluminaGlobal Screening array (Illumina Inc, San Diego, CA) at the Erasmus MC, The Netherlands. Before imputation, AT and CG SNPs were removed. Imputation was performed using shapeit2 and impute2. Chromosomal and base pair positions were updated to the 1000 Genomes Phase 3 reference set, allele strands were flipped where necessary. After imputation, we reran quality control, filtering out SNPs with an info score < 0.8, a minor allele frequency below 1% and a deviation from HWE with a p-value < 1.0e-06. This resulted in a final dataset of 6,397,528 SNPs.

#### Placental methylation

The quality control of the DNAm data, including 812,911 DNAm probes, was performed using the PACEAnalysis R package (v.0.1.7 ; https://www.epicenteredresearch.com/ ). With the R package, we discarded those samples with a call rate below 95%, sex inconsistencies, intentioned or non-intentioned duplicates, and those contaminated with DNA from another subject or the mother (discarded samples N = 23).

Only those samples with paired genotype data were considered in this study. Probes with a call rate lower than 95% and in the sex chromosomes were excluded from the analysis.

The methylation beta values were normalized in different steps. Dye-bias and Noob background correction, implemented in the minfi R package, were applied, followed by normalization of the data with the functional normalization method. Then, to correct for the bias of type-2 probe values, the beta-mixture quantile (BMIQ) normalization was applied. After that, we explored the clustering of the data through Principal Component Analysis (PCA) and tested the association of the 12 first PCs with the main and the technical variables. Array batch effect was controlled with the ComBat method. To correct for the possible outliers, we Winsorized the extreme values to the 1% percentile (0.5% on each side), where percentiles were estimated with the empirical beta distribution. Finally, the rank-based inverse normal transformation (RNT) was applied to the beta values, and these estimations were the DNAm values considered for mQTL mapping. The final dataset consisted of 117 samples and 748 879 DNAm probes (CpGs).

Cell type proportions of six populations (STB, TB, nucleated red blood cells, Hofbauer cells, endothelial cells, and stromal cells) were estimated from DNAm using the placenta reference panel from the 3rd trimester implemented in the Planet R package.

**Placental *cis*-mQTL analysis**

A total of 6,397,528 SNPs and 748 879 CpGs from 117 samples with paired genotype and DNAm data were considered for the *cis*-mQTL analysis in TensorQTL. TensorQTL nominal modality performs linear regressions between the genotype and the normalized DNAm RNT values, as implemented in FastQTL. The *cis*-region was specified as ±0.5Mb from each tested CpG position.

The covariates included in the regression model were the sex of the fetuses, the first five PCs derived from the genotype data (genotype PCs), the first 5 non-genetic DNAm PCs, and the cell type proportions calculated with the Planet methylation panel. Genotype and DNAm PCs were included in the model as covariates to remove hidden and/or technical confounders affecting the DNAm data. To avoid multicollinearity between the non-genetic DNAm PCs and the other covariates, the DNAm PCs were calculated on the residuals from a multiple linear regression adjusting the normalized DNAm RNT-values by the known covariates (sex of the fetuses, the first five genotype PCs, and the five estimated cell types). Following Min et al., we kept all DNAm PCs that cumulatively explained 80% of the variance with a maximum number of 20 PCs for subsequent steps. Then we performed a GWAS on each of the DNAm PCs and retained those PCs that were not associated with the genotype at a suggestive threshold ($P > 1 \times 10^{-7}$). This procedure returned 5 nongenetic DNAm PCs.

**Funding**

The PREDO Study has been funded by the Academy of Finland (J.L.: 311617 and 269925, K.R.: 1312670 ja 128789 1287891), EraNet Neuron, EVO (a special state subsidy for health science research), University of Helsinki Research Funds, the Signe and Ane Gyllenberg foundation, the Emil Aaltonen Foundation, the Finnish Medical Foundation, the Jane and Aatos Erkko Foundation, the Novo Nordisk Foundation, the Päivikki and Sakari Sohlberg Foundation, Juho Vainio foundation, Yrjö Jahnsson foundation, The Finnish Society of Sciences and Letters, Jalmari and Rauha Ahokas foundation, Sigrid Juselius Foundation granted to members of the Predo study board. Methylation assays were funded by the Academy of Finland (269925). Dr. Lahti has received research support from the Strategic Research Council (SRC) established within the Academy of Finland (decision number: 352700).

**Acknowledgements**

The PREDO study would not have been possible without the dedicated contribution of the PREDO study group members: E Hamäläinen, E Kajantie, H Laivuori, PM Villa, A-K Pesonen, A Aitokallio-Tallberg, A-M Henry, VK Hiilesmaa, T Karipohja, R Meri, S Sainio, T Saisto, S Suomalainen-Konig, V-M Ulander, T Vaitilo (Department of Obstetrics and Gynaecology, University of Helsinki and Helsinki University Central Hospital, Helsinki, Finland), L Keski-Nisula, Maija-Riitta Orden (Kuopio University Hospital, Kuopio Finland), E Koistinen, T Walle, R Solja (Northern Karelia Central Hospital, Joensuu, Finland), M Kurkinen (Päijät-Häme Central Hospital, Lahti, Finland), P.Taipale. P Staven (Iisalmi Hospital, Iisalmi, Finland), J Uotila (Tampere University Hospital, Tampere, Finland). We thank all the PREDO children and their parents for their enthusiastic participation. We also thank all the research nurses, research assistants, and laboratory personnel involved in the Predo study.

**References**

Chen et al., Epigenetics. 2013 Feb 1; 8(2): 203–209. Discovery of cross-reactive probes and polymorphic CpGs in the Illumina Infinium HumanMethylation450 microarray

McCartney et al., Genom Data. 2016 Sep; 9: 22–24.,Identification of polymorphic and off-target probe binding sites on the Illumina Infinium MethylationEPIC BeadChip

Price et al, Epigenetics Chromatin. 2013 Mar 3;6(1):4. Additional annotation enhances potential for biologically-relevant analysis of the Illumina Infinium HumanMethylation450 BeadChip array.

### Rhode Island Child Health Study (RICHS)

**Design and study population**

The Rhode Island Child Health Study (RICHS) is a study of mother-infant pairs with non-pathologic pregnancies that were enrolled from the Women and Infants' Hospital in Providence, RI, USA between September 2010 and February 2013. Mothers younger than 18 years of age, with life threatening conditions, pregnancies resulting in preterm birth (< 37 weeks gestation), or congenital/chromosomal abnormalities were excluded. Study protocols were approved by the institutional review boards (IRB) at the Women and Infants Hospital of Rhode Island and Emory University and all participants provided written informed consent. Infants born small for gestational age (≤ 10$^{th}$ BW percentile) or large for gestational age (≥ 90$^{th}$ BW percentile) were oversampled, then infants adequate for gestational age (between the 10th and 90th BW percentiles) that were matched on gestational age and maternal age were coincidentally enrolled. Sociodemographic, lifestyle and clinical data were collected via study questionnaires and structured medical record abstraction.

**Placental biopsies and DNA extraction**

RICHS collected placental tissues (within 2 hours of delivery) biopsied from the fetal side adjacent to the cord insertion site after removing maternal decidua. Samples were placed in RNAlater (Life Technologies, Carlsbad, CA) then frozen at -80°C. DNA was extracted (Norgen Biotek, Thorold, ON) and quantified via the Qubit Flourometer (Life Technologies), then subsequently stored at -80°C.

**Genotype data**

A subset of placenta DNA samples from the RICHS study were genotyped on the Illumina Expanded Multi-Ethnic Genotyping Array (Mega-EX) (dbGaP accession phs001586.v1.p1). The quality control steps included removing SNPs with low call rate (<0.9), markers deviating from Hardy-Weinberg equilibrium ($P < 1x10^{-6}$) and rare SNPs (< less 5 minor allele count). 1,730,225 SNPs passed quality control and were used for genotype imputation using the Haplotype Reference Consortium as reference panel (version r1.1 2016) [4]. This panel contains individuals with European ancestry and the 1000 Genomes Project data. In total, 5,748,854 SNPs of high imputation quality and minor allele count (MAC).

**DNA methylation data**

The EZ Methylation kit (Zymo Research, Irvine, CA) was used for bisulfite modification. DNA methylation was measured on the Illumina Infinium HumanMethylation450K BeadArray (Illumina, San Diego, CA) at the University of Minnesota Genomics Center. Samples were randomized across batches; batch variables were recorded to allow for corrections. Raw array data are available via the NCBI Gene Expression Omnibus (GEO) accession number GSE75248. Array image files (IDAT) preprocessing and data quality control was performed in R using the PACEAnalysis R package (v.0.1.7) that includes background correction, detection and exclusion of poor-quality probes (p-value < 0.01), assessment of concordance between array-inferred sex and phenotypic sex, and exclusion of contaminated samples using the SNPs in the array. We used the ComBat method to array batch effects [13]. DNA methylation Beta values were normalized with probe type dye bias adjustment (BMIQ), Noob background correction followed the functional normalization (minfi R package [31]). We winsorized extreme values to the 1% percentile (0.5% on each side), where percentiles were estimated with the empirical beta distribution. Finally, the rank-based inverse normal transformation (RNT) was applied to beta values, and these were used for mQTL mapping. The final dataset consisted of 167 samples and 423,141 DNAm CpGs. With the DNA methylation array data, we estimated six placental cell type proportions (syncytiotrophoblast, trophoblast, nucleated red blood cells, Hofbauer cells, endothelial cells, and stromal cells) using the 3rd trimester placenta reference from the Planet R package [15].

**Placental gene expression data**

RNA was isolated with RNeasy Mini Kit (Qiagen, Valencia, CA), quantified via Nanodrop Spectrophotometer (Thermo Scientific, Waltham, MA), and stored at -80°C. Placental RNA-seq data from a subset of samples (n = 200) were obtained using the Illumina Hi-Seq 2500 platform. Approximately 20 million single-end RNA-seq reads were generated on each sample. Quality control was performed with FastQC. Sequencing reads were aligned to the human reference genome using STAR and summarized with featureCounts. DSeq2 R package and normalized using the variance-stabilizing transformation. Raw reads are available at the NCBI sequence read archive (SRP095910).

**Placental *cis*-mQTL analysis**

A total of 5,277,034 SNPs and 423,141 CpGs from 167 samples with paired genotype and DNAm data were considered for the *cis*-mQTL analysis in TensorQTL. TensorQTL nominal modality performs linear regressions between the genotype and the normalized DNAm RNT values, as implemented in FastQTL. The *cis*-region was specified as ±0.5Mb from each tested CpG position.

The covariates included in the regression model were the sex of the fetuses, the first five PCs derived from the genotype data (genotype PCs), the first 17 non-genetic DNAm PCs, and the cell type proportions calculated with the Planet methylation panel. Genotype and DNAm PCs were included in the model as covariates to remove hidden and/or technical confounders affecting the DNAm data. To avoid multicollinearity between the non-genetic DNAm PCs and the other covariates, the DNAm PCs were calculated on the residuals from a multiple linear regression adjusting the normalized DNAm RNT-values by the known covariates (sex of the fetuses, the first five genotype PCs, and the five estimated cell types). Following Min et al., we kept all DNAm PCs that cumulatively explained 80% of the variance with a maximum number of 20 PCs for subsequent steps. Then we performed a GWAS on each of the DNAm PCs and retained those PCs that were not associated with the genotype at a suggestive threshold ($P > 1 \times 10^{-7}$). This procedure returned 17 nongenetic DNAm PCs.

**Placenta eQTM analysis**

Placental DNAm and RNA-seq data were used for expression quantitative trait methylation analysis (n=195). eQTMs were calculated by implementing linear models in MatrixEQTL, considering a cis window of 0.5 Mb up and downstream of each CpG. Linear regressions were adjusted by sex, five PCs of expression, and the Planet estimated cell types. Results were corrected with FDR.

**Acknowledgement**

We thank all the RICHS participants and their families.

**Funding**

The RICHS study was supported by the National Institutes of Health (NIH) with the following grant: R01MH094609.

### Bibliography


1. Dadvand, P., et al., *Cohort Profile: Barcelona Life Study Cohort (BiSC).* Int J Epidemiol, 2024. **53**(3).
2. Purcell, S., et al., *PLINK: a tool set for whole-genome association and population-based linkage analyses.* Am J Hum Genet, 2007. **81**(3): p. 559–75.
3. Jin, Y., et al., *GRAF-pop: A Fast Distance-Based Method To Infer Subject Ancestry from Multiple Genotype Datasets Without Principal Components Analysis.* G3 (Bethesda), 2019. **9**(8): p. 2447–2461.
4. McCarthy, S., et al., *A reference panel of 64,976 haplotypes for genotype imputation.* Nat Genet, 2016. **48**(10): p. 1279–83.
5. Campagna, M.P., et al., *Epigenome-wide association studies: current knowledge, strategies and recommendations.* Clin Epigenetics, 2021. **13**(1): p. 214.
6. Binder, A.M. *PACE Analyses*. Available from: https://www.epicenteredresearch.com/pace/.
7. Vespalcova, H., et al., *Association of in utero exposure to phthalate and DINCH metabolites with placental DNA methylation.* (1873-6750 (Electronic)).
8. Zhou, W., et al., *SeSAMe: reducing artifactual detection of DNA methylation by Infinium BeadChips in genomic deletions.* Nucleic Acids Res, 2018. **46**(20): p. e123.
9. Heiss, J.A. and A.C. Just, *Identifying mislabeled and contaminated DNA methylation microarray data: an extended quality control toolset with examples from GEO.* Clin Epigenetics, 2018. **10**: p. 73.
10. Fortin, J.P., T.J. Triche, Jr., and K.D. Hansen, *Preprocessing, normalization and integration of the Illumina HumanMethylationEPIC array with minfi.* Bioinformatics, 2017. **33**(4): p. 558–560.
11. Triche, T.J., Jr., et al., *Low-level processing of Illumina Infinium DNA Methylation BeadArrays.* Nucleic Acids Res, 2013. **41**(7): p. e90.
12. Fortin, J.P., et al., *Functional normalization of 450k methylation array data improves replication in large cancer studies.* Genome Biol, 2014. **15**(12): p. 503.
13. Teschendorff, A.E., et al., *A beta-mixture quantile normalization method for correcting probe design bias in Illumina Infinium 450 k DNA methylation data.* Bioinformatics, 2013. **29**(2): p. 189–96.
14. Johnson, W.E., C. Li, and A. Rabinovic, *Adjusting batch effects in microarray expression data using empirical Bayes methods.* Biostatistics, 2007. **8**(1): p. 118–27.
15. Yuan, V., et al., *Cell-specific characterization of the placental methylome.* BMC Genomics, 2021. **22**(1): p. 6.
16. Min, J.L., et al., *Genomic and phenotypic insights from an atlas of genetic effects on DNA methylation.* Nat Genet, 2021. **53**(9): p. 1311–1321.
17. Newschaffer, C., et al., *The EARLI Study as a Resource for Research on Autism and the Environment.* Epidemiology, 2012. **23**(5S).
18. Heude, B., et al., *Cohort Profile: The EDEN mother-child cohort on the prenatal and early postnatal determinants of child health and development.* (1464-3685 (Electronic)).
19. Anderson, C.A., et al., *Data quality control in genetic case-control association studies.* Nat Protoc, 2010. **5**(9): p. 1564–73.
20. Taylor, S., et al., *A practical approach to RT-qPCR-Publishing data that conform to the MIQE guidelines.* Methods, 2010. **50**(4): p. S1–5.
21. Bourgey, M., et al., *GenPipes: an open-source framework for distributed and scalable genomic analyses.* Gigascience, 2019. **8**(6).

22. Li, H. and R. Durbin, *Fast and accurate long-read alignment with Burrows-Wheeler transform.* Bioinformatics, 2010. **26**(5): p. 589–95.
23. McKenna, A., et al., *The Genome Analysis Toolkit: a MapReduce framework for analyzing next-generation DNA sequencing data.* Genome Res, 2010. **20**(9): p. 1297–303.
24. DePristo, M.A., et al., *A framework for variation discovery and genotyping using next-generation DNA sequencing data.* Nat Genet, 2011. **43**(5): p. 491–8.
25. Institute, B. *Picard*. 2025; Available from: http://broadinstitute.github.io/picard/.
26. Rubinacci, S., et al., *Efficient phasing and imputation of low-coverage sequencing data using large reference panels.* Nat Genet, 2021. **53**(1): p. 120–126.
27. Byrska-Bishop, M., et al., *High-coverage whole-genome sequencing of the expanded 1000 Genomes Project cohort including 602 trios.* Cell, 2022. **185**(18): p. 3426–3440.e19.
28. Sherry, S.T., et al., *dbSNP: the NCBI database of genetic variation.* Nucleic Acids Res, 2001. **29**(1): p. 308–11.
29. Guxens, M., et al., *Cohort Profile: the INMA-INfancia y Medio Ambiente-(Environment and Childhood) Project.* Int J Epidemiol, 2012. **41**(4): p. 930–40.
30. Hertz-Picciotto, I., et al., *A Prospective Study of Environmental Exposures and Early Biomarkers in Autism Spectrum Disorder: Design, Protocols, and Preliminary Data from the MARBLES Study.* Environ Health Perspect, 2018. **126**(11): p. 117004.
31. Aryee, M.J., et al., *Minfi: a flexible and comprehensive Bioconductor package for the analysis of Infinium DNA methylation microarrays.* Bioinformatics, 2014. **30**(10): p. 1363–9.
